# The Impact of Schools on the Transmission of Sars-Cov-2: Evidence from Italy^*^

**DOI:** 10.1101/2022.07.18.22276940

**Authors:** Salvatore Lattanzio

## Abstract

This paper studies the effect of school re-openings and closures on the spread of Sars-Cov-2 in Italy. Exploiting different re-opening dates across regions after the summer break 2020, I show that early opening regions experience on average 1,900 more cases per day in the 40 days following school re-openings compared with late opening ones. However, the uncertainty around the estimates is large and suggests a wide dispersion in the effects of school re-openings on Sars-Cov-2 transmission. I also study the effect of school closures in Campania, one of the biggest regions in Southern Italy. Using a synthetic control approach, I show that school closures are associated with lower case numbers relative to the counterfactual group, particularly in younger age groups. In contrast, I find no significant effects on older age groups, who are more likely to require hospitalization. Finally, exploiting survey data on incidence rates in schools, I provide descriptive evidence on the increased incidence among teachers and students relative to the general population, following school re-openings.

## 1. Introduction

School closures are a widely used measure to contain the spread of COVID-19. According to the Oxford COVID-19 Government Response Tracker,^1^ all 185 countries present in their database adopted some form of school closure to contain the pandemic from January 2020. The rationale behind school closures is that reduced contacts among students and mixing in small spaces should limit the diffusion of COVID-19. However, in the presence of a virus that implies highly heterogeneous likelihoods of developing severe symptoms for young and old individuals, as Sars-Cov-2, school closures can backfire if students meet outside of schools among themselves and with older age groups (e.g., with grandparents and relatives with caring responsibilities). In this case, school closures can ease transmission among more vulnerable individuals, causing additional pressure on the healthcare system. Hence, whether schools contribute to the circulation of the virus is an empirical question.

The literature has investigated the effectiveness of school closures to contain the spread of other respiratory viruses in the past. For example, Jackson et al. (2013) provide a review of the evidence on the effects of school closures and re-openings on pandemic and seasonal influenza outbreaks. They suggest that closing schools is effective in curbing the infection rate, whereas school reopenings tend to be associated with higher viral transmission. Adda (2016) reports similar results, finding that school closures play a significant role in reducing the spread of viral diseases. However, the paper suggests that closing schools for extended periods of time is not cost-effective. Based on the knowledge available at the time, policymakers opted for school closures in the wake of the Sars-Cov-2 pandemic. In fact, model-based evidence highlights that the impact of schools on the transmission of the virus is relevant, but depends on the amount of out-of-school interactions among children (Head et al. 2022). A variety of empirical contributions have investigated the impact of school closures and school openings on the transmission of COVID-19, reaching different conclusions depending on the context, the time of the year when the study was conducted, and the stringency of the other non-pharmaceutical interventions that were in place. Different research suggests that schools either play a role in the increase of COVID-19 cases, or they do not. Studies done in Germany, Sweden, and Japan suggest that there is no correlation between schools reopening and an increase in cases (Isphording, Lipfert, and Pestel 2021; von Bismarck-Osten, Borusyak, and Schonberg 2022; Vlachos, Hertegård, and B. Svaleryd 2021; Fukumoto, McClean, and Nakagawa 2021). However, research done in the United States suggests the opposite, that schools are in fact playing a role in the increase of COVID-19 cases (Goldhaber et al. 2021; Chernozhukov, Kasahara, and Schrimpf 2021; Courtemanche et al. 2021). Moreover, Bravata et al. (2021) provide convincing evidence that transmission increases in households where school-age children are present.

This paper contributes to the literature by providing evidence on both the effects of school re-openings and closures on the diffusion of COVID-19, using Italy as a case study. The focus on Italy is justified by the availability of two quasi-experimental settings connected to policymakers’ decisions on school openings and closures. First, school re-openings after the summer break were staggered across regions and, in 2020, two groups of regions opened schools on 14 and 24 September, allowing a comparison of infections in the two sets. Second, local authorities were allowed, during the pandemic, to adopt policy measures that were more restrictive than those mandated at national level, comprising school closures. Therefore, I exploit school closures in Campania—one of the biggest Southern regions—on 16 October and investigate if they had a significant effect in curbing infections. I conduct these analyses using publicly available regional-level data on infections, hospitalizations and deaths provided by the Italian *Protezione Civile* and incidence rates by age groups collected by the Italian Association of Epidemiologists. In both cases, I find a significant, yet imprecise, effect of schools on infection rates. Early re-opening of schools led to a surge in cases and hospitalizations relative to late opening regions. The point estimates suggest that early opening regions would have experienced approximately 1,900 less cases per day over a 40-day horizon, had they reopened schools later (around 47 percent of daily cases experienced in the same period). These estimates should nonetheless be taken with caution as they are characterized by wide uncertainty. On the other hand, school closures in Campania led to a marginally significant drop in incidence rates relative to a synthetic matched group of regions that did not close schools until later in the year. The effect of school closures on reduced Sars-Cov-2 circulation is, however, concentrated in younger age groups, while the effects on older individuals are not statistically significant. Hence, school closures seem ineffective in shielding the age groups that are more vulnerable to develop severe symptoms of COVID-19.

Finally, using data collected by the Ministry of Education, I show that—in contrast with existing evidence (e.g., Gandini et al. 2021)—incidence in schools, especially among the teaching and non-teaching staff and students aged 14-18, is higher than that in the general population in the period during which schools were open.

This paper therefore contributes to the growing literature that investigates the impact of schools on Sars-Cov-2 transmission and provides evidence on a country, Italy, that was severely hit by both first and second waves in Spring and Autumn 2020, and that extensively used school closures as one of the main non-pharmaceutical interventions to curb infection rates. The closest paper to mine in the Italian context is Amodio et al. (2022) who, using detailed geo-localized data on infection rates and variation in school openings time in September 2020 for Sicily, find that census areas where schools opened early witness an increase in COVID-19 cases by 1.5-2.9 percent. This paper provides additional evidence by focusing on the whole country and by investigating the impact of both school openings and closures, as they may have non-symmetric effects on infections. My results are in line with studies on the United States (Courtemanche et al. 2021), but are in contrast with evidence from other European countries, e.g., Germany (Isphording, Lipfert, and Pestel 2021; von Bismarck-Osten, Borusyak, and Schonberg 2022), where school openings did not contribute to a higher viral transmission. Differences between Italy and Germany may be due to a variety of factors. Social interactions between children and compliance with mask mandates within schools, the behavior of parents, public transport use and quality, aeration within classrooms, and the structure and type of school buildings, are all possible explanations of why schools may have a different impact in the two countries. To shed light on this point, I investigate how crowding in classrooms and the age of school buildings are associated with infections among students in different points in time, providing some descriptive evidence that both play a role.

In general, heterogeneous results in different countries can depend on the stringency of non-pharmaceutical interventions in place. As highlighted, if schools are closed but students meet outside with less precautions than those that schools would mandate, then school closures can be quite ineffective in reducing the circulation of the virus. Moreover, the effectiveness of school closures further depends on the strain of the virus circulating in the community, with more contagious variants (e.g., Delta and Omicron) being additional elements of risk; on population immunity, which in turn depends on past infection levels and vaccine coverage; and on policy measures adopted to make schools safer, as the use of N95 masks or the availability of air filtering systems. Therefore, the divergence in findings in the literature highlights even more the importance of studying the impact of schools on Sars-Cov-2 transmission in different contexts and time periods to verify the external validity of the results.^2^

It is important to highlight that I only focus on the impact of schools on infections, while ignoring the potential negative short- and long-run effects of school closures on learning. The costs of school closures for students can be high and persistent, especially if they have low socio-economic background. Agostinelli et al. (2022) highlight how online learning is an imperfect substitute for in-person schooling and that lost interactions between students, combined with differential investment of parents from high socio-economic background in their children’s education, contribute to persistent learning losses. Halloran et al. (2021) show that, in the United States, there was a substantially larger decline in pass rates on standardized tests in Grades 3-8 in districts with remote schooling compared to those with in-person schooling, with more severe losses in districts with larger populations of Black students. Similarly, Contini et al. (2022) show that school closures had a negative effect on students’ achievement in mathematics, using data from Italy. Engzell, Frey, and Verhagen (2021), and Fuchs-Schündeln et al. (2020) highlight how school closures are likely to widen gaps in learning and future career outcomes between children from high- and low-income families. Parolin and Lee (2021) further show that, in the US, school closures increased inequality, as they were more likely to happen in schools with higher shares of students who are from ethnic minorities, have experienced homelessness, and are eligible for free or reduced-price lunch. Finally, school closures can have negative effects on children’s mental health (Lee 2020) and affect disproportionately women, who are usually demanded a higher share of childcare within households (Alon et al. 2020).^3^ Given these potential negative effects, it is even more important to provide convincing answers on the effects of schools on Sars-Cov-2 transmission.

## 2. Sars-Cov-2 in Italy and Schools

Italy was the first Western country to be hit severely by COVID-19. The first case dates back to 31 January, whereas the first death was registered on 21 February. Following the diffusion of the virus, especially in the North of the country, the government opted for the implementation of two “red zones”, involving 11 municipalities in Lombardy and Veneto, that were effectively in lock-down. At the same time, many Northern regions opted to close schools. This measure was then extended to the whole nation on 4 March, just a few days before the nation-wide lockdown, which was established on 10 March. Schools remained closed throughout the whole school year and only high-school students taking the final exam were allowed in classrooms during the summer. In September schools reopened in different days in different groups of regions. Figure 1 shows the opening dates of schools in each Italian region. There are two main blocks: those that opened on 14 September and those that opened on 24 September. The former are mainly concentrated in the Centre-North (with the exception of Molise and Sicily), whereas the latter are concentrated in the South of the country. Three regions opened schools in different days: Bolzano on 7 September, Friuli Venezia Giulia on 16 September and Sardinia on 22 September. In the empirical application, I will focus on regions opening on 14 and 24 September.

**Figure 1.**
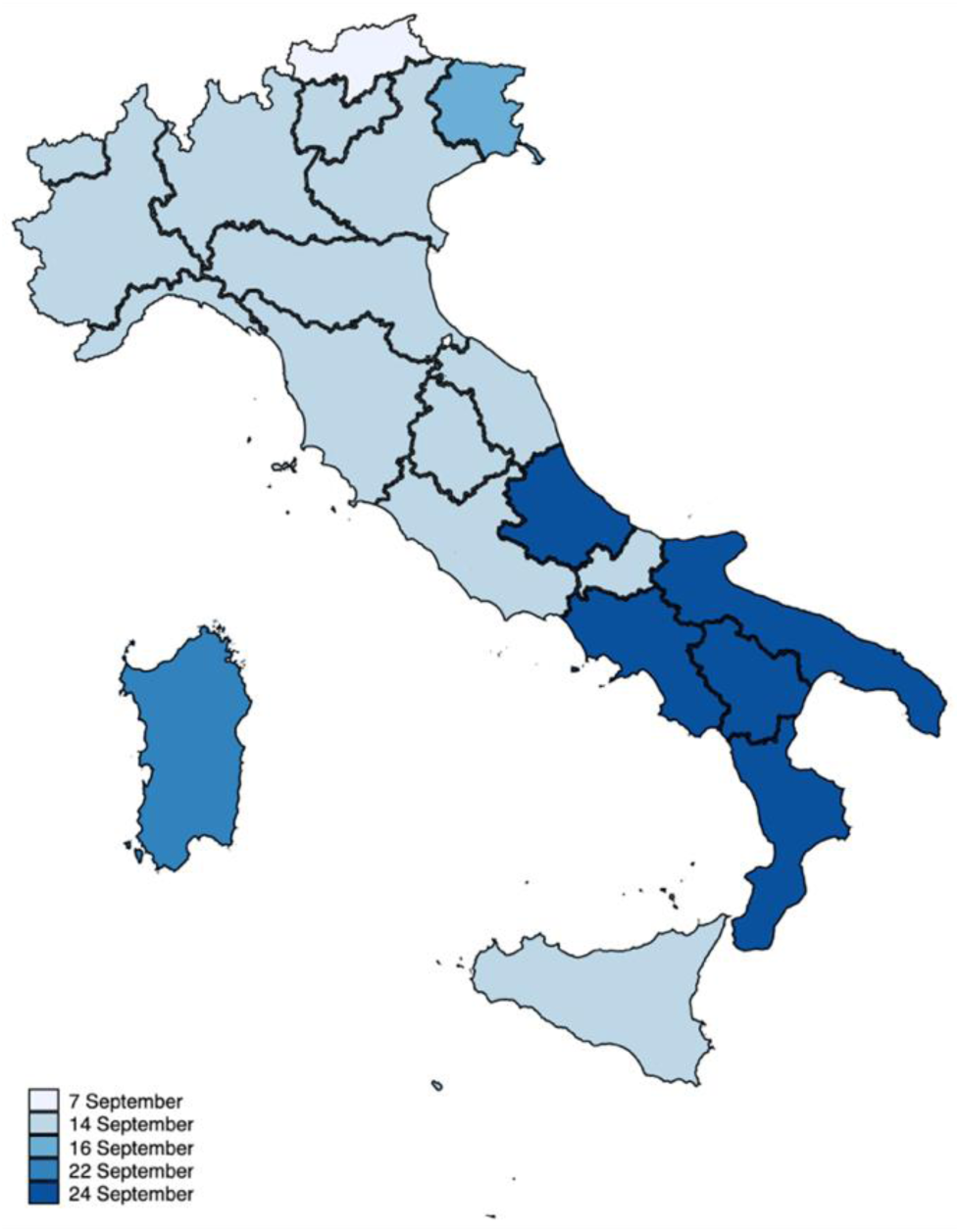
School opening dates after the summer in Italy *Notes*. The figure displays a map of Italy with the administrative boundaries of the 19 regions and two autonomous provinces. Different colors indicate different opening dates of schools after the summer break.

Following school re-openings case numbers remained low throughout September and started rising in October in a second wave that then led the government to establish a range of non-pharmaceutical interventions that comprised partial or total school closures depending on the incidence of the disease. In anticipation of government decisions, Campania opted to close all schools on 16 October to curb infection rates. This decision was unexpected and taken in isolation by the president of the region and was not followed by other regions, which decided instead to follow government advice. On November 4, the government introduced new local lockdown measures, depending on the level of circulation of the virus in each region. These measures comprised partial school closures, depending on the stringency of measures adopted. In particular, the government established three different areas of risk, based on three colors: yellow, orange and red. In yellow and orange zones, high schools (and universities) were closed, whereas kindergarten, elementary and middle schools remained open. In red zones, only kindergarten, elementary schools and the first year of middle schools were operating in-person.

## 3. Data and Descriptive Statistics

### Data

I combine several data sources. First, I exploit data on the number of new cases, hospitalizations, patients in intensive care units and deaths provided by the Italian *Protezione Civile*. The data provide daily figures from the end of February 2020 for each Italian region and, for provinces,^4^ they provide the total number of cases since the beginning of the pandemic. I compute per capita quantities using data on population—as of January 2020—from the national statistical institute (Istat). Second, I exploit information on weekly incidence rates by age group provided by the Italian Association of Epidemiologists, which makes the data publicly available for 13 regions starting from 21 September.^5^ Third, I use data from the Ministry of Education on the number of cases in schools in students aged 6-13 and 14-18 and among the teaching and non-teaching staff, which are publicly available from the study of Gandini et al. (2021).^6^ Fourth, I use data on mobility from Google Reports to control for mobility differences across regions, that could affect the diffusion of the virus. Fifth, I use data on the characteristics of municipalities from Istat to construct a matched control group and study the impact of school closures in Campania: average family size, the number of students or residents who use public transports to go to school or work per 100,000 residents, the number of university students per 100,000 residents, population density and the share of population between 60 and 79 years old and over 80. Finally, I use data from the Ministry of Education^7^ on the number of students per square meter and on the age of all schools in each Italian region in order to explore possible associations between incidence among students and school buildings’ quality or classroom crowding.

### Descriptive Statistics

Figure 2 reports the regional evolution of the number of new cases per 100,000 residents in September and October 2020. The figure suggests that, on average, where schools opened earlier cases increased at a faster pace relative to regions opening later, although the dispersion of the time series widens considerably over time.

**Figure 2.**
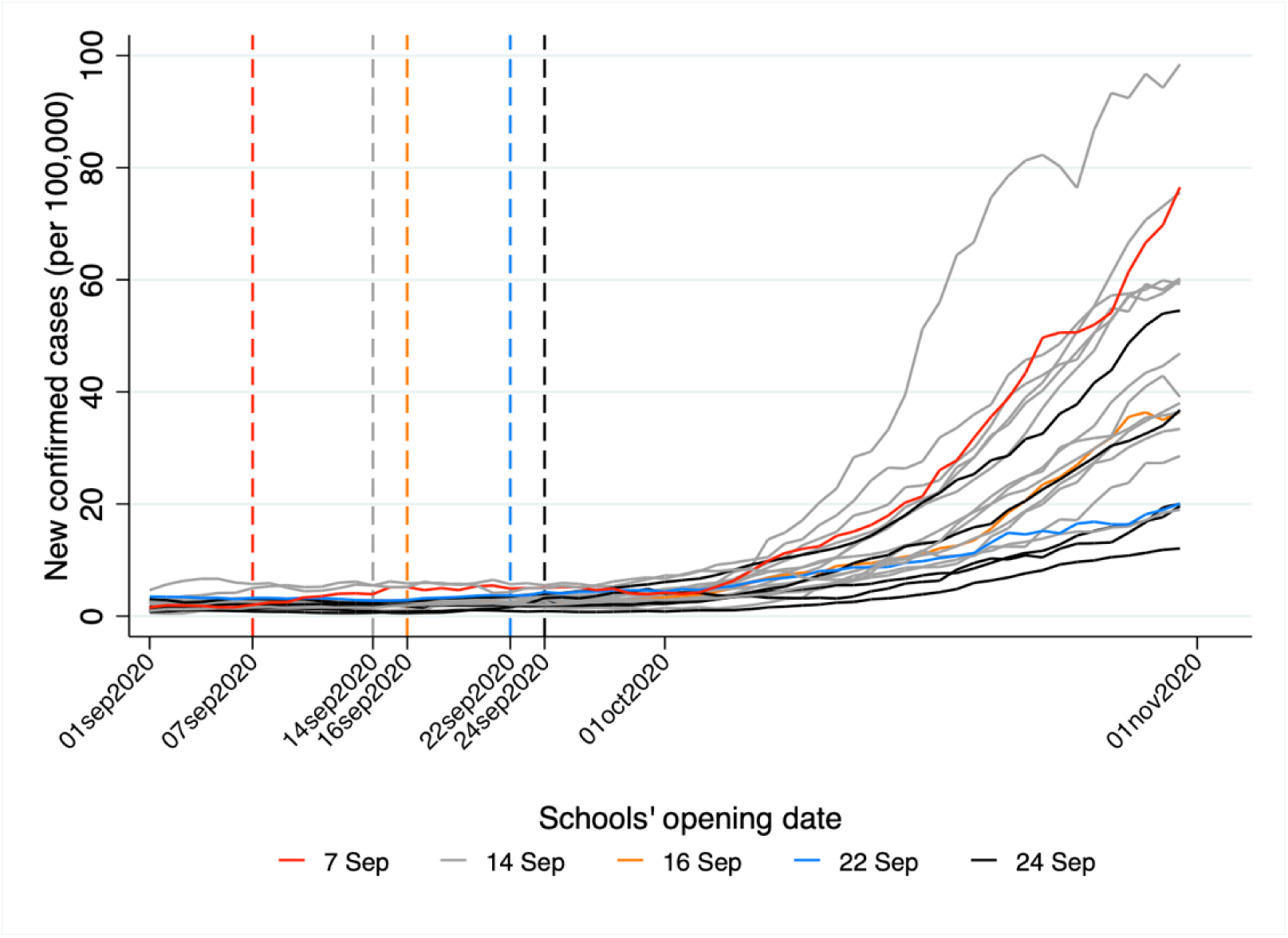
New cases per 100,000 residents by region and time around school re-openings in September *Notes*. The figure shows the evolution of daily cases per 100,000 residents in each region. Different colors indicate different opening dates of schools. Vertical dashed lines are put in correspondence of each opening date. Source: author’s own calculation based on data from *Protezione Civile*.

How was mobility affected by school re-openings? Figure 3 displays the evolution of mobility relative to the reference period of January 2020 from Google Mobility Reports in September and October 2020, separately for regions opening schools on 14 and 24 September (respectively, early and late opening regions). The figure reports average changes in mobility by week^8^ between 30 August and 2 November relative to a baseline period—i.e., the median value, for the corresponding day of the week, during the period 3 January-6 February, 2020. The figure shows that mobility towards transit stations displays diverging patterns between the two groups of regions in the moments when they re-opened schools—although it is generally below the baseline level—and then decreases from October onward, probably because of classrooms in quarantine or because students avoided public transports to reach schools. Other mobility categories display comparable evolutions over time. Mobility towards retailing decreases, following school openings, in both groups of regions. Residential mobility increases relative to the baseline following school openings, probably because of schools offering hybrid teaching systems with part of lectures being in presence and part at home. Mobility towards workplaces is below the baseline in both early and late opening regions. At the end of October, when cases began to increase exponentially, mobility to workplaces decreases, as a consequence of an increased use of remote working.

**Figure 3.**
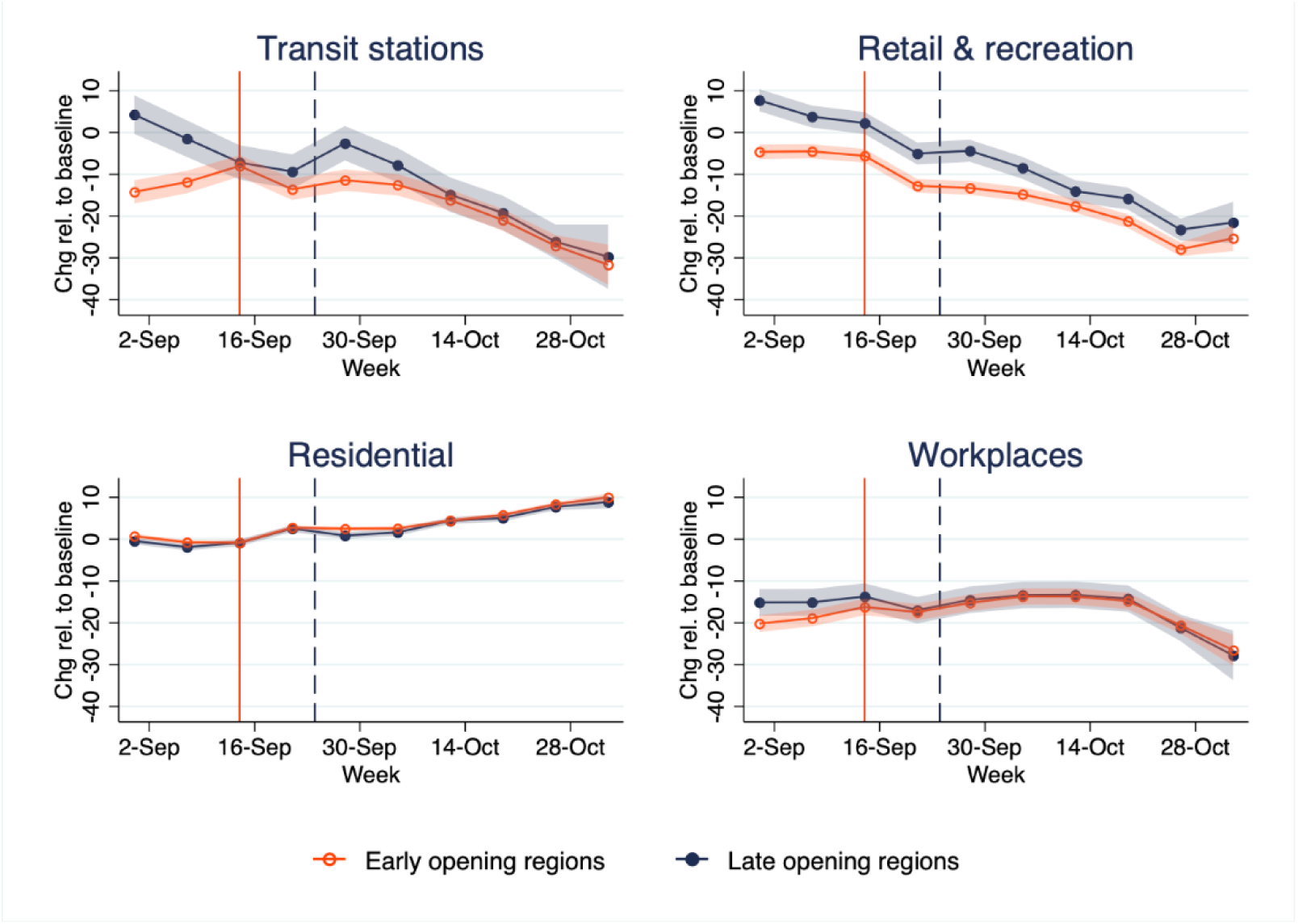
Mobility trends in September and October 2020 *Notes*. The figure reports mobility trends relative to a baseline period, i.e., the median value, for the corresponding day of the week, during the period 3 January-6 February, 2020. Each dot in the graph is the predicted value from a regression of mobility on week dummies, treatment dummy and the interaction of both. 95 percent confidence intervals are recovered from standard errors obtained via the delta method. Source: author’s own calculations based on Google Mobility Reports.

## 4. Empirical Strategy

### School re-openings

I focus on regions that opened schools on 14 or 24 September and limit the observation window to the period of time between 15 August and 3 November. I therefore include a long pre-treatment period of 30 days and a post-treatment period of 40 days. I do not go beyond 3 November, as on that day the government issued new restrictive measures that affected schools. On this sample, I estimate the following difference-in-differences model:

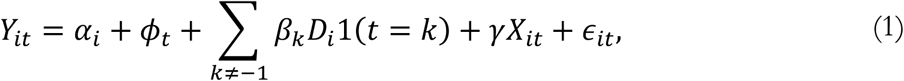

where *Y*_*it*_ is the number of new positive cases, daily change in hospitalizations and in the number of patients in intensive care units, or daily deaths. *α*_*i*_ are region fixed effects that control for time-invariant unobserved factors, such as the age composition of the population and population density. *ϕ*_*t*_ are calendar date fixed effects controlling for unobserved shocks that are common across regions (e.g., nation-wide interventions). *D*_*i*_ is a dummy variable equal to one for regions opening schools on 14 September and zero for those opening on 24 September. 1(*t* = *k*) are event time dummies, using the day before re-openings (i.e., 13 September) as a reference point. *X*_*it*_ includes region-level controls for mobility towards grocery and pharmacy, parks, retail and recreational places, workplaces, and homes.^9^ Finally, *ϵ*_*it*_ is an error term. The coefficients of interest are the *β*_*k*_’s which measure the daily change in outcomes between early and late opening regions relative to the day before re-openings.

### School closures in Campania

To evaluate the impact of school closures in Campania, I use a synthetic control method and compare the evolution in the number of daily cases per 100,000 residents and in the incidence rates by age group in Campania and in a synthetic group of regions that act as control units. I then report the average outcomes for treated and synthetic units and their difference for each period *t* ∈ {1, …, *T*}, where time is either days or weeks:

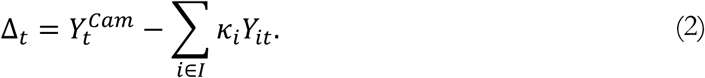

In (2), *i* indexes regions and *κ*_*i*_ are the weights assigned to each region. I limit the observation window to the week starting on 21 September until the one starting on 30 November. There are two “treatment periods”: one in which Campania is *fully* treated, between 16 October and 3 November; one in which Campania is *partially* treated, between 4 and 30 November. The fully treated period corresponds to the days in which only Campania closed schools, whereas the partially treated period corresponds to days in which, following nation-wide policy measures, also other regions closed some school orders, depending on the stringency of measures adopted. In low-risk areas (depending on the circulation of the virus in the community), high schools (and universities) were closed, whereas kindergarten, elementary and middle schools remained open. In high-risk areas, only kindergarten, elementary schools and the first year of middle schools were operating in-person. In both *fully* and *partially* treated periods, all school orders were closed in Campania. The variables used to create the synthetic control group are population, population density, the number of individuals using public transports for work- and school-related reasons per 100,000 residents, the number of cases in the first wave (21 February-3 May) per 100,000 residents, the share of residents between 60 and 79 years old and older than 80, the number of university students per 100,000 residents and the average number of family components. Moreover, when analyzing total cases, I also include the change in daily hospitalizations and in the number of patients in ICUs, the daily number of deaths, and the tests performed per 100,000 residents. When analyzing incidence rates by age group, I further include incidence rates for the adjacent younger and two older age groups. For example, when computing the synthetic control for the age group 25-44, I include incidence rates for the group 19-24, 45-59 and 60-69 as matching variables.^10^ Table A1 reports weights applied to each region for the synthetic control for each different outcome.

## 5. Results

### 5.1. The Impact of School Re-openings After the Summer

I start by investigating the impact of school re-openings in September. Figure 4 reports the estimates of *β*_*k*_ from equation (1). Panel (a) shows results for daily new positive cases. Before school re-openings, regions that opened early were on parallel trends with respect to late opening regions. After schools opened, there are no statistically significant differences between the two groups of regions for approximately 25 days. However, after 25 days, differences begin to emerge, which become larger and statistically significant between the 30^th^ and 40^th^ day following re-openings on 14 September. On average, early opening regions witnessed an increase of 10.1 additional new daily cases with respect to late opening regions between 25 and 40 days after re-openings. Panel (b) reports coefficients for the daily change in hospitalizations, which shows a similar, although less precise, pattern. Differences between early and late opening regions are not statistically significant before re-openings and in the 25 days following them. After 25 days, regions opening schools early witness an increase of 0.7 hospitalizations per 100,000 residents. Panel (c) shows that there are no significant differences in terms of the number of patients in intensive care units both before and after school openings. Panel (d) reports results for daily deaths: although there is a spike at the very end of the observation window, differences between treated and control units are never statistically significant at conventional levels. The absence of differences in the number of critically ill patients and deaths can be interpreted as a good sign if one believes that most of the increase in cases in early opening regions happen among younger subjects, who are less exposed to the risk of developing severe symptoms from COVID-19. However, the time horizon may be too short to observe differences in the number of critically ill patients and deaths, given the lag between symptoms onset and more severe clinical developments of the disease. It would be however difficult to go beyond the observation window reported in the graphs, which ends on 3 November, as from that date there are other non-pharmaceutical interventions adopted by the government that could confound the effects.

**Figure 4.**
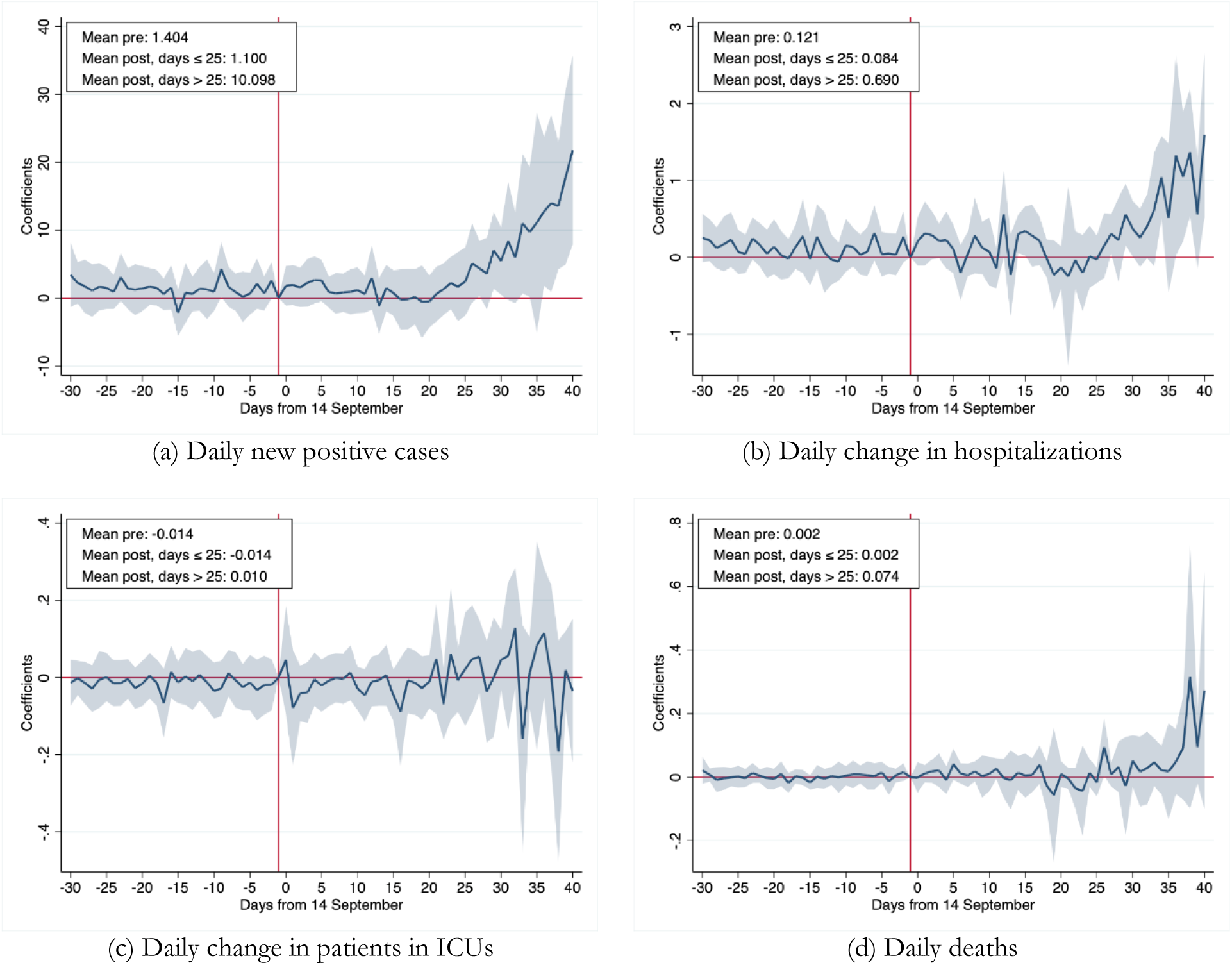
Difference-in-differences coefficients for cases, hospitalizations, ICUs and deaths for regions opening early vs late around school openings *Notes*. The figure reports estimates of dynamic coefficients *β*_*k*_ from equation (1). All outcomes are values per 100,000 residents. The shaded areas are 95 percent confidence intervals obtained from cluster-robust standard errors at the region level.

In order to assess the magnitude of the results, I use the estimates from the regressions for daily cases to predict how cases would have evolved if schools had reopened later in early opening regions. Specifically, I compute the sum of point estimates in the 40 days following school reopenings,^11^ which implies that the cumulative number of extra daily cases in early opening regions is 180.1 per 100,000 residents. The total number of extra cases can be obtained by multiplying 180.1 by the early opening regions’ total population of 42,884,254 and dividing by 100,000, yielding 77,229 or an average of 1931 daily cases. In the 40 days following school re-openings there were a total of 164,372 new cases in the early opening regions or an average of 4109 daily cases. Hence, this back-of-the-envelope calculation suggests that early re-opening regions would have experienced 47 percent lower case numbers had schools re-opened 10 days later. These numbers must be interpreted with caution, as they display wide uncertainty. They are not statistically significant: the 95 percent confidence interval ranges between -18 and 112 percent. Focusing, instead, on the period 9-24 October (i.e., from the 26^th^ to the 40^th^ day following early re-openings), early opening regions would have experienced 4184 daily cases less, had schools opened later, i.e., as much as 50 percent of average daily cases in that period (95 percent confidence interval: 2-99 percent).

The increase in the number of cases is not a consequence of increased testing capacity of early opening regions, as Figure A1 in the Appendix shows. Panel (a) reports estimates of equation (1) using the daily number of tests administered per 100,000 residents as outcome and shows that there are no significant differences between the two groups of regions. Panel (b) reports results for the test positivity rate, computed as the ratio between daily cases and tests, and shows that there is a marked increase in the positivity rate 25 days after re-openings.

One concern with these results is that, except for Molise and Sicily, early opening regions are concentrated in the Centre and North of the country. Although all regressions include region fixed effects—therefore controlling for time-invariant characteristics of the two groups—and mobility controls, there might still be time varying unobserved confounders that drive the results. Therefore, I re-run equation (1) excluding regions in the North. I further exclude Campania, as the region closed schools on 16 October: a decision that, as Section 5.2 details, has an impact on the circulation of the virus. The resulting sample includes eight regions: four in the treated group (Lazio, Molise, Sicilia, Umbria) and four in the control group (Abruzzo, Basilicata, Calabria, Puglia). Figure A2 in the appendix shows the estimates. Overall, the results are broadly confirmed if one focuses on the number of daily cases. The estimates become less precise, though. There is nonetheless a clear increasing trend and the difference amounts to 8.3 additional daily cases per 100,000 residents in regions opening schools early. In contrast with the findings in the specification with all regions, hospitalizations remain flat following re-openings. Finally, the figure confirms that there is no effect on patients in intensive care units and deaths.

As a further robustness check, Figure A3 reports coefficients using the daily change in total cases (which sums cases, hospitalizations, and deaths) at the province level for the full sample of regions in panel (a) and the restricted sample of regions in the Centre-South in panel (b). The patterns at the province level are similar to those at the region level, with little differences emerging between provinces in early and late opening regions within three to four weeks since school re-openings and a significant difference showing up approximately after 25 days.

One potential source of bias in the estimates comes from elections held on 20-21 September. On these dates, all citizens voted for a referendum to reduce the number of members of Parliament. This referendum does not represent a threat to identification, as it was held in all regions at the same time and was not preceded by electoral campaigns that brought to large gatherings of people.

However, seven regions (Campania, Liguria, Marche, Puglia, Toscana, Valle d’Aosta and Veneto) also voted to renew their presidents. Cipullo and Le Moglie (2022) show that the electoral campaigns preceding regional elections influenced the circulation of the virus:^12^ where local elections were held, the number of cases, hospitalizations and deaths is significantly larger than in places without local elections, as large gatherings of people may have occurred in the former group during electoral campaigns. It is not easy to control for the impact of elections in equation (1). Any indicator for the presence of local elections or for voter turnout would be absorbed by region fixed effects. Reassuringly, regional elections are held in 5 out of 13 regions in the treated group of early opening regions and in 2 out of 5 regions in the control group of late opening regions. Hence, remarkably similar shares of 38 and 40 percent of both groups hold regional elections. Furthermore, I re-run equation (1) excluding regions that held elections. Figure A4 reports the estimates of the dynamic difference-in-differences coefficients for daily cases. Panel (a) reports results for the sample that includes regions in the North and in the Centre-South, whereas panel (b) focuses on regions in the Centre-South only.^13^ Both figures display patterns that are similar to those in the main analysis. The increase in daily cases, 25 days after re-openings, in early opening regions is lower than that in the preferred sample: 6.7 vs. 10.1 new daily cases per 100,000 residents when focusing on the entire country, and 7.1 vs. 8.3 when focusing on regions in the Centre-South only. These more conservative estimates provide a lower bound to the effect of schools on Sars-Cov-2 transmission, though differences between different samples are not statistically significant.

### 5.2. The Impact of School Closures in Campania

I now turn to the inspection of school closures in Campania. Figure 5 reports the 7-day moving average^14^ of new daily positive cases in Campania and in the synthetic control group of regions in panel (a), and the difference between Campania and the synthetic control in panel (b). In both graphs, the first vertical line is 16 October, i.e., the day when schools closed in Campania. The second vertical dashed line is 3 November, i.e., the day when the government adopted a new set of non-pharmaceutical interventions. I report the evolution of cases until the end of November, therefore comparing outcomes over the *fully treated* period (16 October-3 November) and the *partially treated* period (4 November-30 November), when also other regions closed schools partially, following nation-wide policy measures.

**Figure 5.**
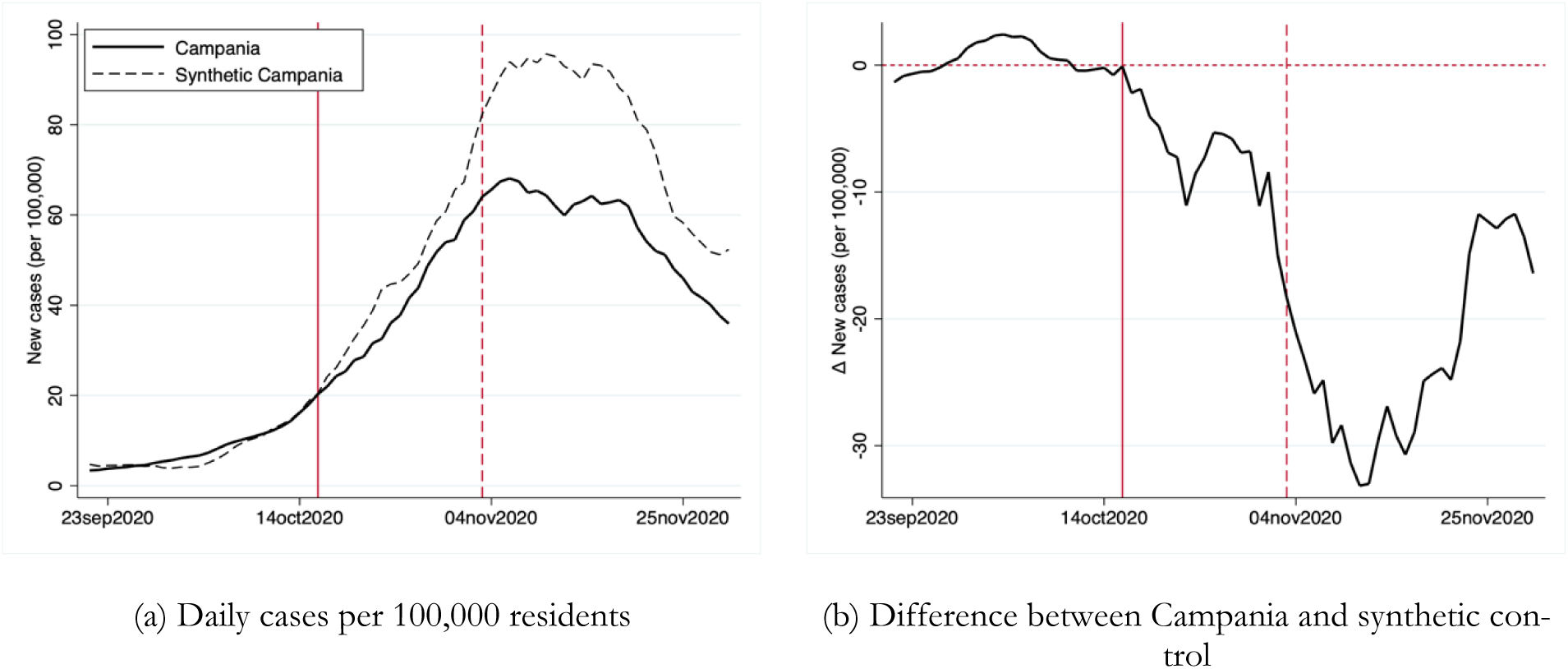
Evolution of new daily cases (7-day moving average) per 100,000 residents in Campania and in the synthetic control group *Notes*. The figure reports in panel (a) the evolution of daily cases in Campania (solid line) and in the synthetic control group (dashed line) between 21 September and 30 November. Panel (b) reports the difference between the two lines. The solid vertical line corresponds to 16 October, when schools were closed in Campania. The dashed vertical line corresponds to 3 November, when new nation-wide non pharmaceutical interventions were adopted by the central government.

The figure shows that before school closures the number of daily cases in Campania matches that of the synthetic control. After school closures in Campania a difference emerges between the two lines: the cumulative number of cases per 100,000 residents in the period 16 October-3 November is 764 in Campania against 902 in the synthetic group, a 15 percent reduction. After 3 November differences between the two groups increase until mid-November, although schools were partially closed also in other regions. However, on the one hand, it may take time for school closures to have an effect on case numbers: hence, the effect observed after 3 November may be a consequence of school closures in the period before. On the other hand, differences may widen between the two groups, because only some types of schools were closed in other regions, whereas *all* of them were closed in Campania.^15^ In the second half of November, differences between Campania and the synthetic control decrease, likely as a consequence of partial lockdown measures adopted in other regions.

The observed differences in the number of daily cases between the two groups may be a consequence of different testing policies. Figure A6 in the appendix shows that, until the last week of October, the evolution of daily tests in Campania and in the synthetic control is broadly comparable. From that moment, and for most of November, Campania conducted *more* tests than the control group. Hence, the lower case numbers in Campania are not attributable to lower testing capacity.

One question is whether these effects are significant also in statistical terms. I follow Abadie, Diamond, and Hainmueller (2010) who propose a falsification test based on the distribution of placebo effects using all units in the control group. The null hypothesis that the effect of school closures in Campania is zero can be rejected if the true estimates are larger in magnitude than the placebo estimates. Figure A7 reports the true estimate (i.e., the difference between the solid and dashed line from Figure 5) and the distribution of placebo estimates using the other regions as treated units. The figure suggests that the estimates for Campania are indeed large compared to most of the placebo estimates over the fully treated period and more so over the partially treated period. If one computes an “empirical p-value” as the number of placebo estimates that are lower in magnitude than the estimate for Campania over the total number of placebo regions (i.e., 20), then the null cannot be rejected at 5 percent level only over the periods 26 October-1 November and 23-27 November.

What age groups were affected by school closures the most? I answer this question by using data on weekly-level incidence rates by age from the Italian Association of Epidemiologists, which reports them for 13 regions (Campania, Emilia-Romagna, Friuli Venezia Giulia, Lazio, Lombardia, Marche, P.A. Trento, Piemonte, Puglia, Sicilia, Toscana, Umbria, Veneto). Figure 6 shows the results. School closures have an impact on the number of cases for children between 6 and 10 years old and between 11 and 13 years old and, to a lesser extent, for children between 3 and 5. The figure also shows the presence of an effect for the age groups 19-24 and 25-44. The former may be affected because Campania, besides schools, closed universities, too, which were then closed in November also in other regions. The latter may comprise parents of school-age children and, therefore, closures may have blocked household transmission. There are no visible effects on older age groups in the full treatment period, whereas there is a reduction in incidence rates for age groups 70-79 and 80-89 in the partial treatment period.

**Figure 6.**
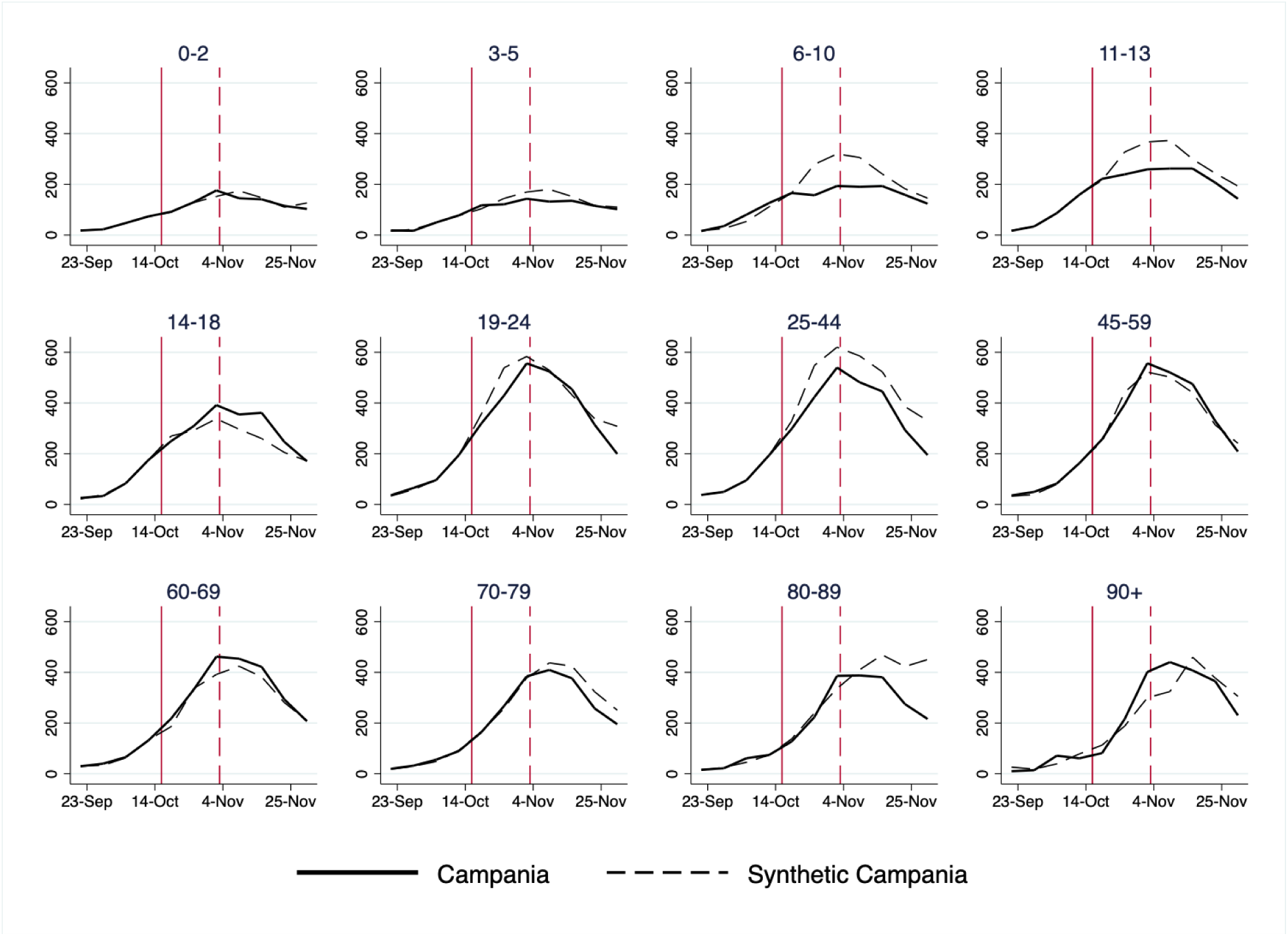
Incidence rates by age group and week in Campania and the synthetic control group *Notes*. The figure reports the evolution of incidence rates by week for different age groups for Campania and the synthetic control. The solid vertical line corresponds to 16 October, when schools were closed in Campania. The dashed vertical line corresponds to 3 November, when new nation-wide non pharmaceutical interventions were adopted by the central government.

It is also interesting to note that school closures do not seem to affect incidence rates for students aged 14-18. It may be the case that high school students can meet outside of schools to a larger extent than younger kids, reducing the effectiveness of school closures. Finally, incidence rates are slightly larger for age groups 60-69 and over 90 in Campania relative to control regions. This may suggest that grandparents have provided additional childcare following school closures increasing their likelihood of being infected.

Figure A8 in the appendix reports the distribution of true and placebo estimates for each age group. For age groups 6-10 and 11-13, estimates are in the lower end of the distribution over the full treatment period, as well as for the age group 25-44. In contrast, the null that school closures had no effect on incidence rates cannot be rejected for older age groups.

Overall, this evidence suggests that school closures had an impact in curbing infections among the younger age groups, but they do not seem to affect, at least in the short run, older age groups, who face a higher risk of being critically ill from COVID-19. In light of this result, the effectiveness of school closures in limiting the pressure on hospitals can be put into question in the presence of viruses that have highly heterogeneous symptoms for young and old individuals. If they have no significant effects on older age groups, school closures may not be enough to reduce cases among those at greater risk of being hospitalized.

### 5.3. A Closer Look at the Pandemic Inside Schools

The question remains open as to whether in-person teaching, or other factors, contribute to rising case numbers. Depending on the answer to this question, policy options may be different. For example, if schools do contribute only through increased congestion on public transports, then the solution would not be to close schools, but rather to increase the frequency and the quality of public transports. The evidence on school closures in Campania, however, suggests that closing schools reduces case numbers in younger age groups, highlighting how spending time in small classrooms may contribute to the spread of the virus. This evidence can be corroborated by another data source, provided by the Ministry of Education, that records the number of positive cases among students, teachers, and the non-teaching staff in schools. These data are collected through surveys filled each week between 19 September and 7 November by school principals. I aggregate the data at the region level and compare the incidence rates among students of primary schools (6-13 years old), high schools (14-18 years old), teachers and non-teaching staff to that of the general population. If the ratio between incidence rates is above 1 it means that the virus circulates more within schools than in the population. Figure 7 reports the relative incidence for each group in all regions over time, highlighting Campania, and for Italy as a whole. The figure shows that, for students in primary schools, aged 6-13, the relative incidence with respect to that of the population is generally below 1 for most regions. However, there is an increasing trend between 3 and 24 October when schools were in-person across the country. Moreover, the relative incidence is computed using the incidence for the *whole* population at the denominator: considering that children usually develop less severe forms of COVID-19 and are more likely to be asymptomatic, it is likely that their relative incidence is underestimated. Hence, especially for younger students relative incidence ratios are to be interpreted as lower bounds of their true values. For high-school students, aged 14-18, the figure shows higher relative incidence, especially in the period between 26 September and 31 October, although with a declining trend, approximately from 17 October. For teachers, we observe a considerably larger incidence rate, especially after 17 October, when for all regions the relative incidence is larger than 1. The non-teaching staff displays a similar pattern. Campania displays a clear declining trend following 17 October, i.e., after school closures. Figure A9 in the Appendix further distinguishes teachers of primary and secondary schools. The patterns in incidence ratios for both groups are broadly similar. Focusing on Italy as a whole, teachers at primary schools have a larger relative incidence, especially after 10 October. Teachers at secondary schools, instead, display lower incidence ratios at the end of the observation window. This evidence, albeit descriptive, suggests that schools contribute to the diffusion of the virus, not only as a by-product of increased congestion on public transports. Moreover, these results are partially in contrast with Gandini et al. (2021) who conclude that schools are not drivers of contagion. However, differently from my analysis, they focus only on the survey of 7 November to compare incidence in schools to that in the general population.

**Figure 7.**
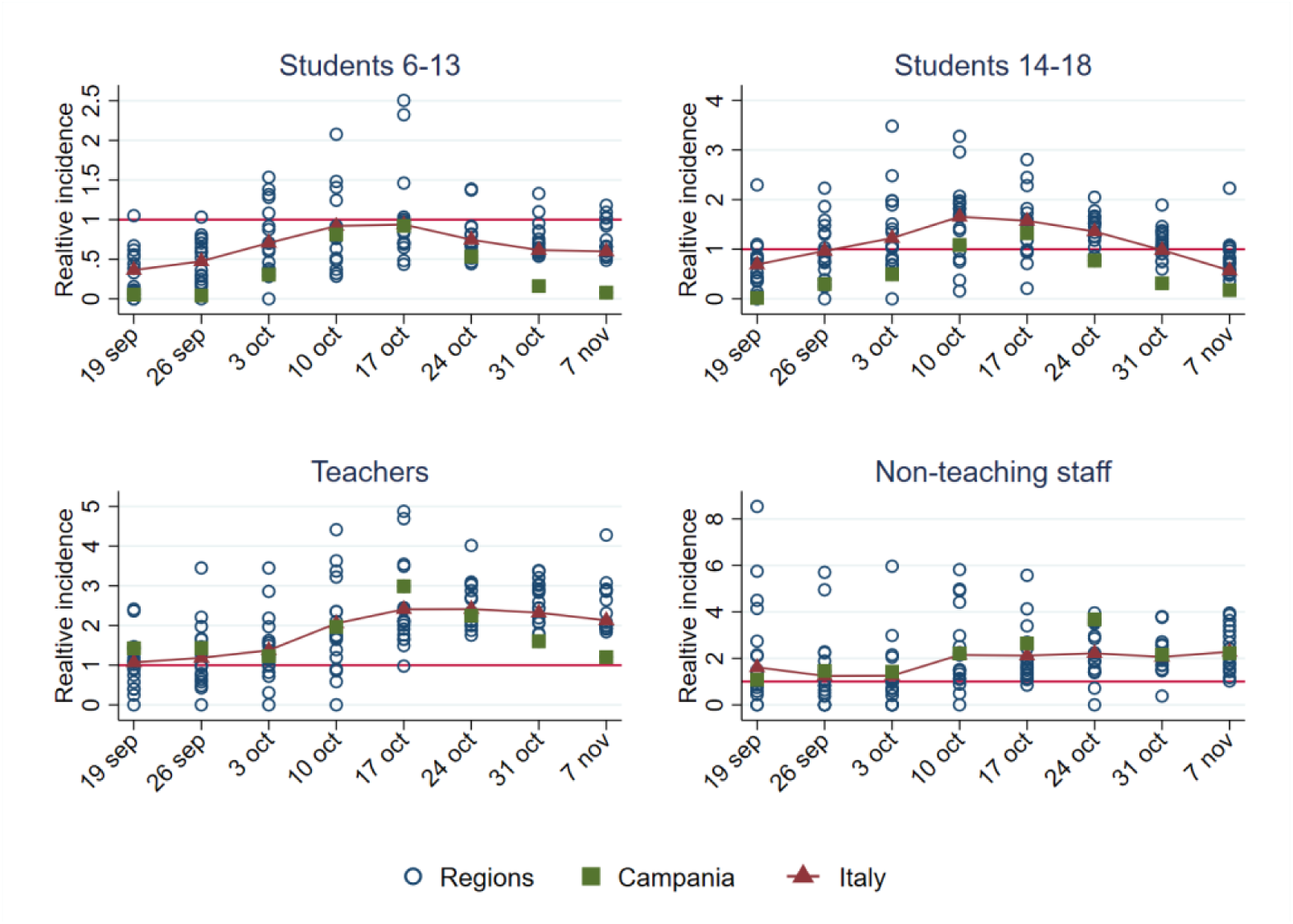
Incidence rate in students, teachers and non-teaching staff relative to that in the general population in Italy and in each region between 19 September and 7 November *Notes*. The figure reports the ratio between incidence in each group (students 6-13, 14-18, teachers, non-teaching staff) and incidence in the general population. Source: author’s own elaboration based on data from the Ministry of Education and *Protezione Civile*.

### 5.4. Mechanisms

What explains the contribution of schools to the transmission of Sars-Cov-2? In this section, I explore two potential mechanisms that could favor the diffusion of the virus in schools. On the one hand, more crowded classrooms can ease contagion as, first, they increase the number of contacts between students and, second, they make it difficult to respect social distancing rules. On the other hand, the age of school buildings can correlate with the spread of the virus, as older buildings may have worse aeration, and smaller spaces and rooms. To test the hypotheses that more crowded classrooms and older buildings correlate with the diffusion of COVID-19, I use data from the Ministry of Education on the number of students per square meter and on the age of buildings for each Italian school. I take the average of the number of students per square meter and the share of schools built before 1976 at the regional level. I then correlate these measures with the relative incidence among students, from section 5.3. Figure 8 reports a scatter plot of the relative incidence, measured in the weeks of 10, 17 and 24 October (around the time of the second wave), against the average number of students per square meter (panel a) and the share of schools built before 1976 (panel b). Panel (a) suggests the presence of a weak positive correlation between the relative incidence among students and classrooms’ crowding. Panel (b) shows the presence of a stronger positive correlation between the relative incidence among students and the share of schools built before 1976, highlighting an association between the spread of the virus and the quality of buildings. In both panels the correlation becomes more evident over time, as the severity of the second wave worsens.

**Figure 8.**
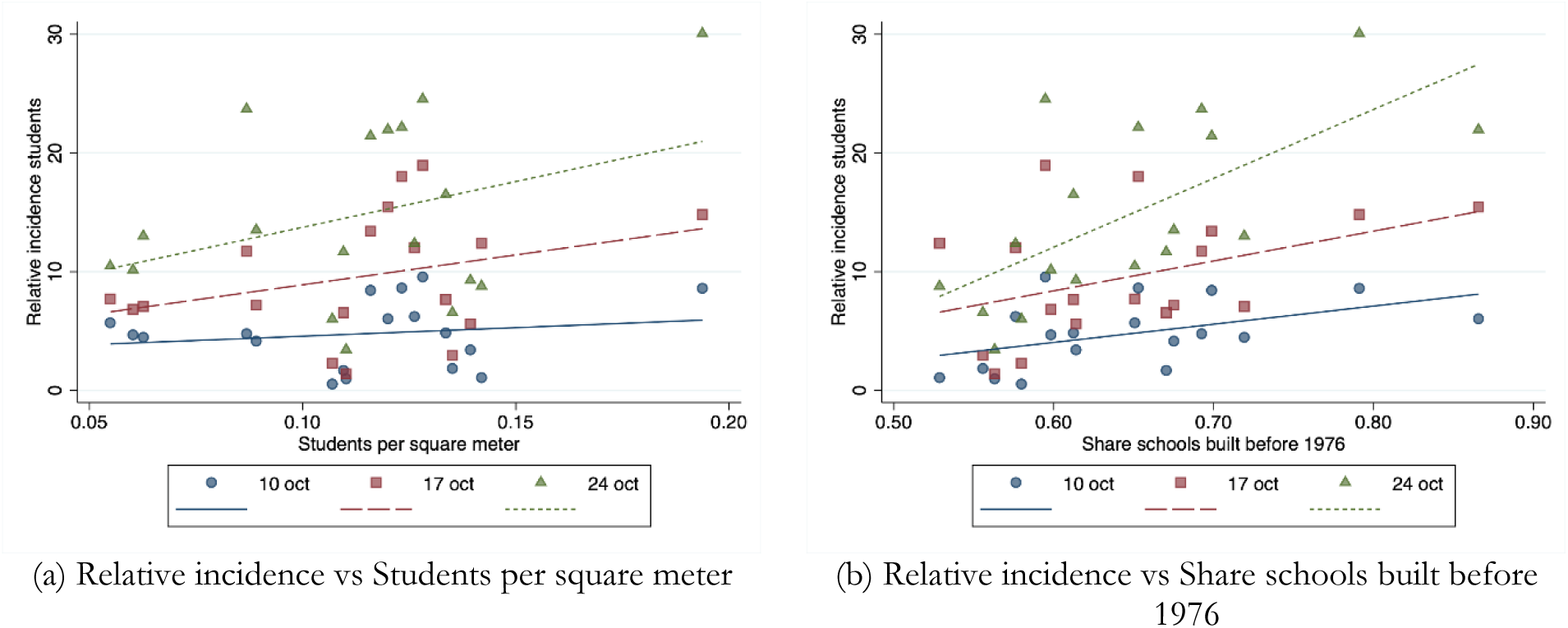
Regional-level scatter plots of the relative incidence among students vs. the number of students per square meter (panel a) and the share of schools built before 1976 (panel b). *Notes*. The figure reports regional level scatter plots and linear fit of the relative incidence among students in the weeks of 10, 17 and 24 October against the number of students per square meter (panel a) and the share of schools built before 1976 (panel b).

Table 1 reports estimates of the association between the incidence among the student population and the number of students per square meter (panel a) and the share of schools built before 1976 (panel b). For each week starting on 10, 17 and 24 October, the table reports estimates without and with controls. Regressions with controls include the region-level GDP per capita in 2019 and the population density to take into account that more prosperous and populated regions have more social and economic interactions, which correlate with the circulation of the virus in the community. The regressions also control for the average level of Particulate Matter 10 (PM10) in each region between 2016 and 2019,^16^ as air pollution has been shown to be associated with higher susceptibility to Sars-Cov-2 (for a review of the evidence, see European Parliament, 2021).

**Table 1.**
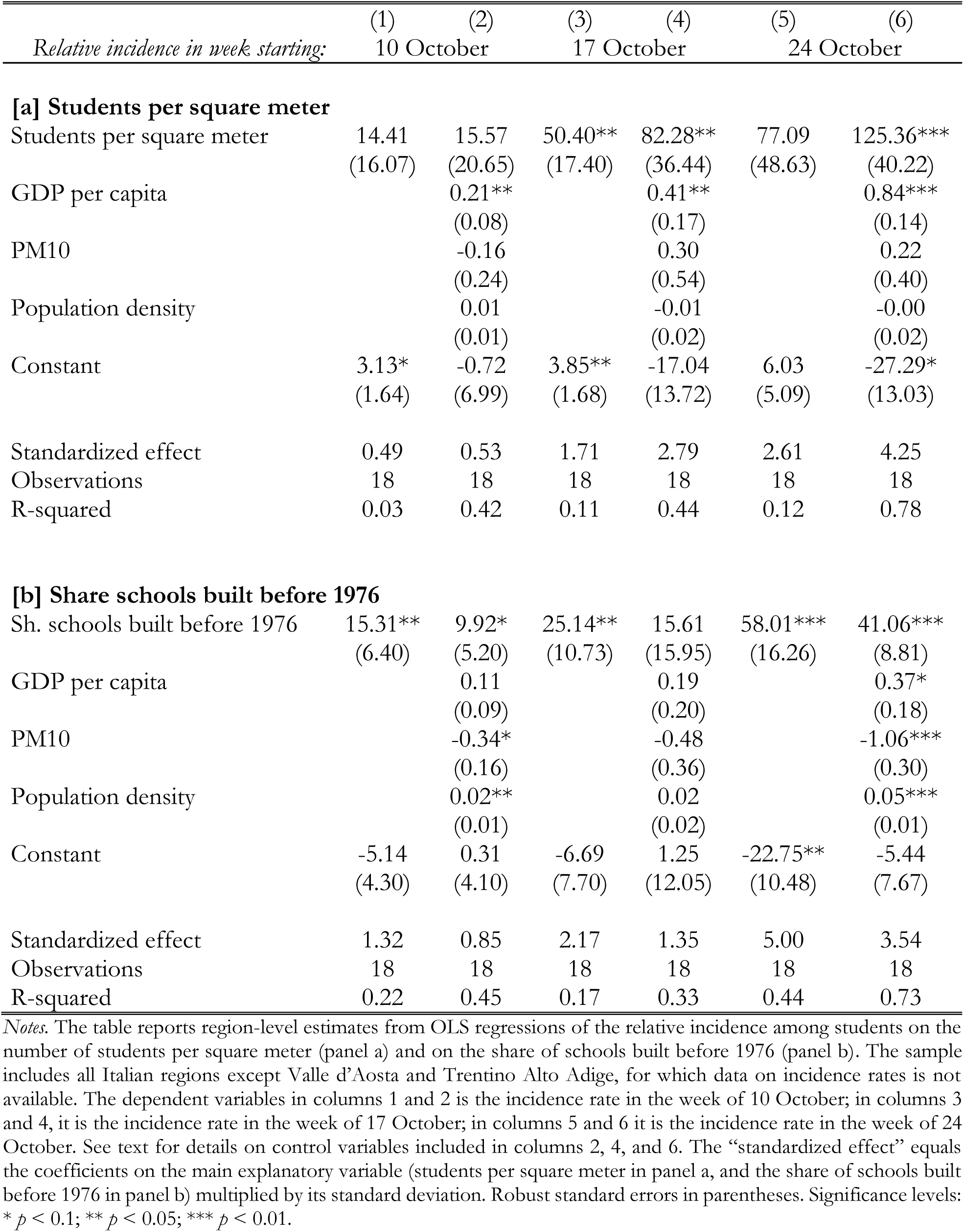
Association between the number of students per square meter and the relative incidence among students, OLS estimates

| *Relative incidence in week starting:* | (1) | (2) | (3) | (4) | (5) | (6) |
| --- | --- | --- | --- | --- | --- | --- |
|  | 10 October |  | 17 October |  | 24 October |  |
| **[a] Students per square meter** |  |  |  |  |  |  |
| Students per square meter | 14.41<br>(16.07) | 15.57<br>(20.65) | 50.40**<br>(17.40) | 82.28**<br>(36.44) | 77.09<br>(48.63) | 125.36***<br>(40.22) |
| GDP per capita |  | 0.21**<br>(0.08) |  | 0.41**<br>(0.17) |  | 0.84***<br>(0.14) |
| PM10 |  | -0.16<br>(0.24) |  | 0.30<br>(0.54) |  | 0.22<br>(0.40) |
| Population density |  | 0.01<br>(0.01) |  | -0.01<br>(0.02) |  | -0.00<br>(0.02) |
| Constant | 3.13*<br>(1.64) | -0.72<br>(6.99) | 3.85**<br>(1.68) | -17.04<br>(13.72) | 6.03<br>(5.09) | -27.29*<br>(13.03) |
| Standardized effect | 0.49 | 0.53 | 1.71 | 2.79 | 2.61 | 4.25 |
| Observations | 18 | 18 | 18 | 18 | 18 | 18 |
| R-squared | 0.03 | 0.42 | 0.11 | 0.44 | 0.12 | 0.78 |
| **[b] Share schools built before 1976** |  |  |  |  |  |  |
| Sh. schools built before 1976 | 15.31**<br>(6.40) | 9.92*<br>(5.20) | 25.14**<br>(10.73) | 15.61<br>(15.95) | 58.01***<br>(16.26) | 41.06***<br>(8.81) |
| GDP per capita |  | 0.11<br>(0.09) |  | 0.19<br>(0.20) |  | 0.37*<br>(0.18) |
| PM10 |  | -0.34*<br>(0.16) |  | -0.48<br>(0.36) |  | -1.06***<br>(0.30) |
| Population density |  | 0.02**<br>(0.01) |  | 0.02<br>(0.02) |  | 0.05***<br>(0.01) |
| Constant | -5.14<br>(4.30) | 0.31<br>(4.10) | -6.69<br>(7.70) | 1.25<br>(12.05) | -22.75**<br>(10.48) | -5.44<br>(7.67) |
| Standardized effect | 1.32 | 0.85 | 2.17 | 1.35 | 5.00 | 3.54 |
| Observations | 18 | 18 | 18 | 18 | 18 | 18 |
| R-squared | 0.22 | 0.45 | 0.17 | 0.33 | 0.44 | 0.73 |

*Notes.* The table reports region-level estimates from OLS regressions of the relative incidence among students on the number of students per square meter (panel a) and on the share of schools built before 1976 (panel b). The sample includes all Italian regions except Valle d'Aosta and Trentino Alto Adige, for which data on incidence rates is not available. The dependent variables in columns 1 and 2 is the incidence rate in the week of 10 October; in columns 3 and 4, it is the incidence rate in the week of 17 October; in columns 5 and 6 it is the incidence rate in the week of 24 October. See text for details on control variables included in columns 2, 4, and 6. The “standardized effect” equals the coefficients on the main explanatory variable (students per square meter in panel a, and the share of schools built before 1976 in panel b) multiplied by its standard deviation. Robust standard errors in parentheses. Significance levels: \* $p < 0.1$ ; \*\* $p < 0.05$ ; \*\*\* $p < 0.01$ .

Panel (a) shows that the correlation between the number of students per square meter and the incidence of cases is positive, but not statistically significant in the week of 10 October (columns 1 and 2), while it becomes statistically significant in the weeks of 17 October (columns 3 and 4) and, when controls are included, in the week of 24 October (column 6). The bottom of the panel reports standardized effect in order to ease the interpretation of the magnitudes: a one standard deviation increase in the number of students per square meter is associated with a 4.3 percent increase in the relative incidence among students in the week of 24 October, in the specification that includes controls (column 6). The aggregate result masks heterogeneity between students in the age groups 6-13 and 14-18. Figure A10 reports separate scatter plots for both age groups in panels (a) and (b), respectively, while Table A2 reports OLS estimates of said relationship. The regression results highlight the presence of a positive correlation for both groups students, albeit noisier for high-school students (panel b). In the week of 24 October, a one standard deviation increase in the number of students between 6 and 13 years old (14 and 18 years old) per square meter increases incidence by 3.2 (5.2) percent.

Panel (b) of Table 1 reports estimates for the share of schools built before 1976 as main explanatory variable. In this case, the correlation is always positive and statistically significant, except when controls are included in the week of 17 October (column 5). Estimates in column (6) suggest that a one standard deviation increase in the region-level share of old schools is associated with a 3.5 percent increase in the relative incidence among students in the weeks of 24 October.

These results, albeit descriptive and based on a small sample, provide a potential explanation of why evidence from different countries reaches different conclusions as to the contribution of schools to the pandemic. Where students have more space available within the school buildings and where the schools are more modern, the diffusion of the virus seems to be slower.^17^

## 6. Conclusion

This paper provides evidence on the interplay between schools and the diffusion of COVID-19. First, it shows that, following re-openings of schools after the summer breaks, early opening regions experience an increase in cases and hospitalizations relative to late opening regions. This is true if one focuses on regions, provinces or on a sub-sample of regions in the Centre-South that are more similar in terms of observable characteristics, although in the latter case hospitalizations display similar trends in early and late opening regions. However, there is wide uncertainty around the estimates. Second, the paper provides evidence on the effectiveness of school closures by exploiting a quasi-natural experiment provided by Campania, where schools of all orders were closed in mid-October, whereas the other regions kept them open. Using a synthetic control, the paper shows that the number of cases decreases in Campania following school closures relative to the control group of regions. The divergence in cases is mainly driven by young children and age groups 19-24 and 25-44. Older age groups are, instead, not affected. Hence, school closures do not seem to influence – at least in the short run – the dynamics of contagion for people who are more exposed to the risk of developing more severe forms of COVID-19. Finally, the paper provides descriptive evidence, using survey data from the Ministry of Education, on the incidence of cases in schools – distinguishing students in primary schools, students in high schools, teachers and the non-teaching staff – relative to that in the general population. This analysis reveals that, especially in October, incidence in schools was significantly higher than that in the general population, although it is not possible to control for differences in testing rates between schools and the rest of the population. Moreover, higher incidence rates among students are associated with the quality of school buildings (measured by their average age at the regional level) and, to a lesser extent, with classrooms’ crowding.

In conclusion, although with some uncertainty around the estimates, schools seem to contribute to the diffusion of the virus. This result is in line with evidence on the United States (Chernozhukov, Kasahara, and Schrimpf 2021; Bravata et al. 2021; Courtemanche et al. 2021), but it is in stark contrast with results on school re-openings in Germany, which show a null effect of school re-openings on Sars-Cov-2 transmission (Isphording, Lipfert, and Pestel 2021; von Bismarck-Osten, Borusyak, and Schonberg 2022). Differences between Italy and Germany may be due to different behaviors of individuals in the two countries, different policy measures to reduce the number of students for each class or to increase air quality within classrooms or different compliance, e.g., with mask mandates within schools. Differences in the quality of public transports may play a role, too. Moreover, the heterogeneity of results in the literature can also depend on the time period being analysed, as different variants of the virus or different degrees of vaccine coverage can affect the relationship between schools and COVID-19. In this sense, it must be noted that all these studies refer to periods in which vaccination was not available: in presence of high immunity levels in the population, schools likely have different effects on the circulation of the virus. It is nonetheless important that more studies become available on the effects of schools on the transmission of Sars-Cov-2 in different countries, as the external validity of findings may be limited and very context-dependent.

Finally, the evidence presented in this paper does not consider the severe and long-lasting effects that school closures have on students’ learning, mental health, social interaction and on parents’— especially mothers’—labour supply. When opting for school closures for a long period of time, policymakers should weigh the benefits of reducing the circulation of the virus against the costs associated with closures. With mass vaccination in the population, the cost-benefit analysis could increasingly be in favor of keeping schools open or, at least, of keeping them open for most of the students on a rotating basis, with hybrid learning systems that combine distance and in-person learning in case of further waves in the future. Having a precise quantification of the benefits of school closures is therefore crucial in order to take appropriate policy responses and avoid unnecessary learning losses for students.

## Data Availability

All data produced in the present study are available upon reasonable request to the authors

## A. Additional Figures and Tables

**Figure A1.**
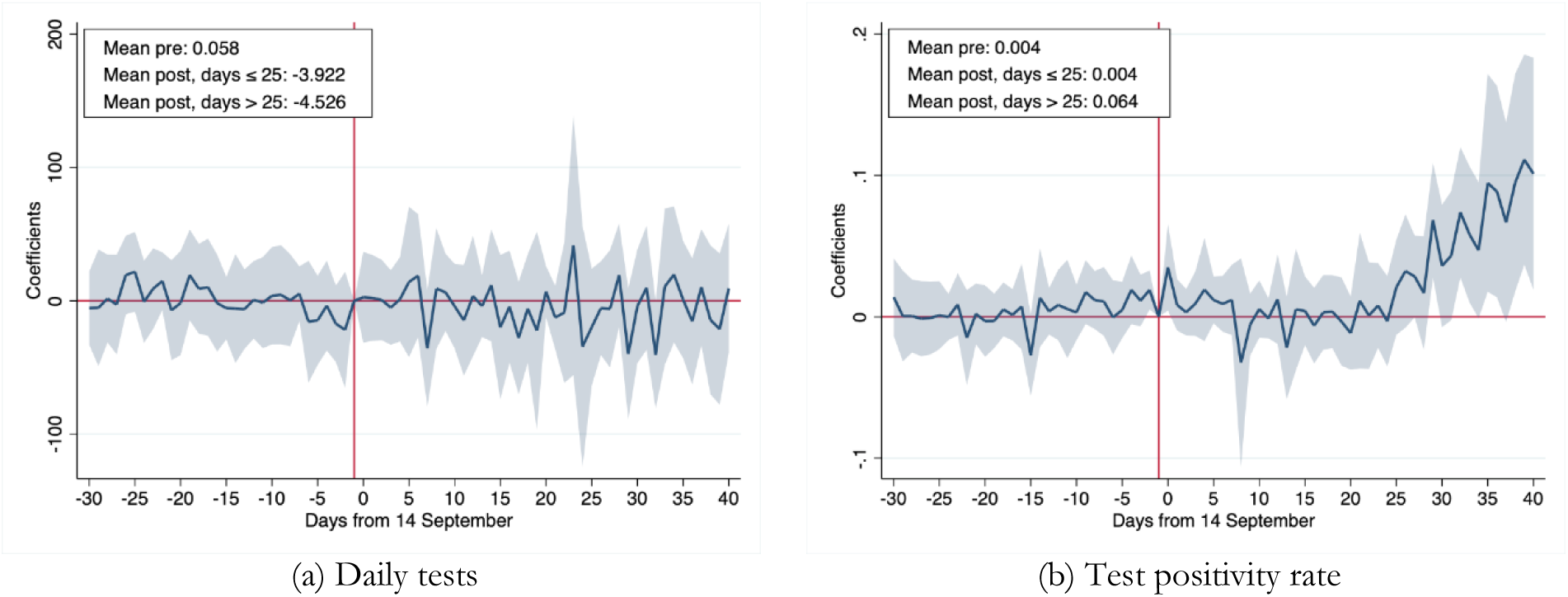
Difference-in-differences coefficients for daily tests and test positivity rate for regions opening early vs late around school openings *Notes*. The figure reports estimates of dynamic coefficients *β*_*k*_ from equation (1). Panel (a) shows results for the number of people tested per 100,000 residents. Panel (b) shows results for the test positivity rate, computed as the ratio of new positive cases over the number of tests performed. The shaded areas are 95 percent confidence intervals obtained from cluster-robust standard errors at the region level.

**Figure A2.**
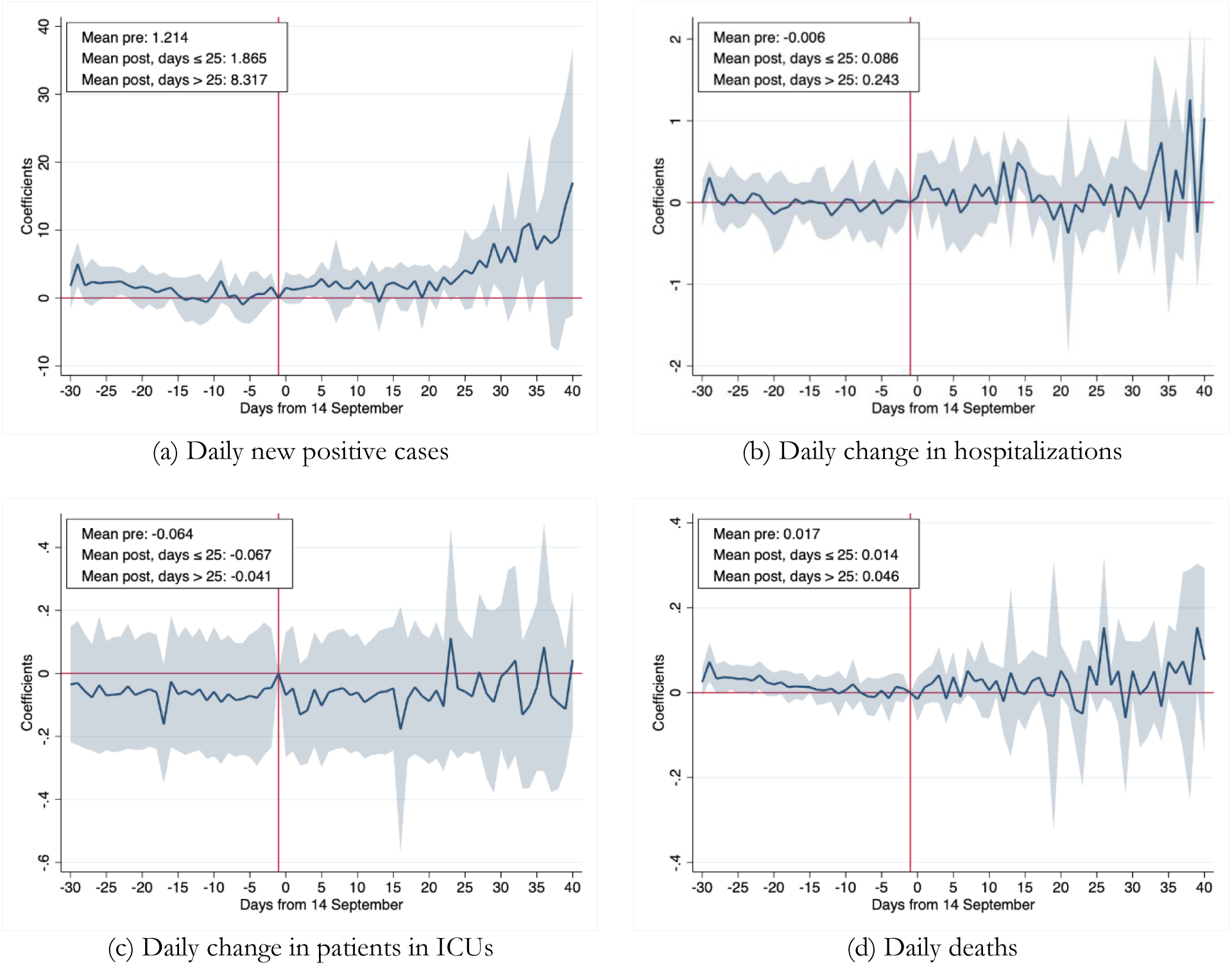
Difference-in-differences coefficients for cases, hospitalizations, ICUs and deaths for regions opening early vs late around school openings, Centre-South regions only. *Notes*. The figure reports estimates of dynamic coefficients *β*_*k*_ from equation (1), restricting the sample to regions in the Centre-South of the country only. Regions included are: Lazio, Molise, Sicilia and Umbria in the treated group; Abruzzo, Basilicata, Calabria and Puglia in the control group. All outcomes are values per 100,000 residents. The shaded areas are 95 percent confidence intervals obtained from cluster-robust standard errors at the region level.

**Figure A3.**
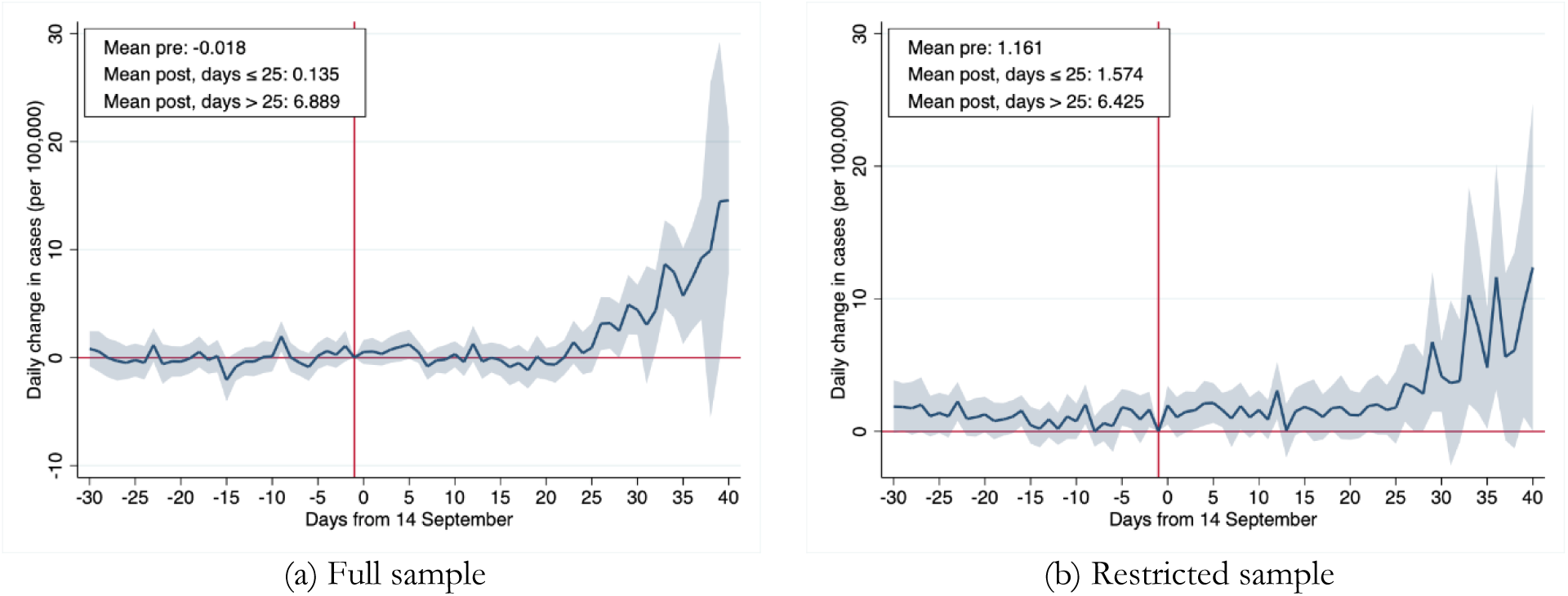
Difference-in-differences coefficients for the daily change in total cases, province level data *Notes*. The figure reports estimates of dynamic coefficients *β*_*k*_ from equation (1) using province-level data on the daily change in total cases. Panel (a) shows results for the full sample of regions. Panel (b) shows results for the restricted sample of regions in the Centre-South: Lazio, Molise, Sicilia and Umbria in the treated group; Abruzzo, Basilicata, Calabria and Puglia in the control group. Outcomes are values per 100,000 residents. The shaded areas are 95 percent confidence intervals obtained from cluster-robust standard errors at the region level.

**Figure A4.**
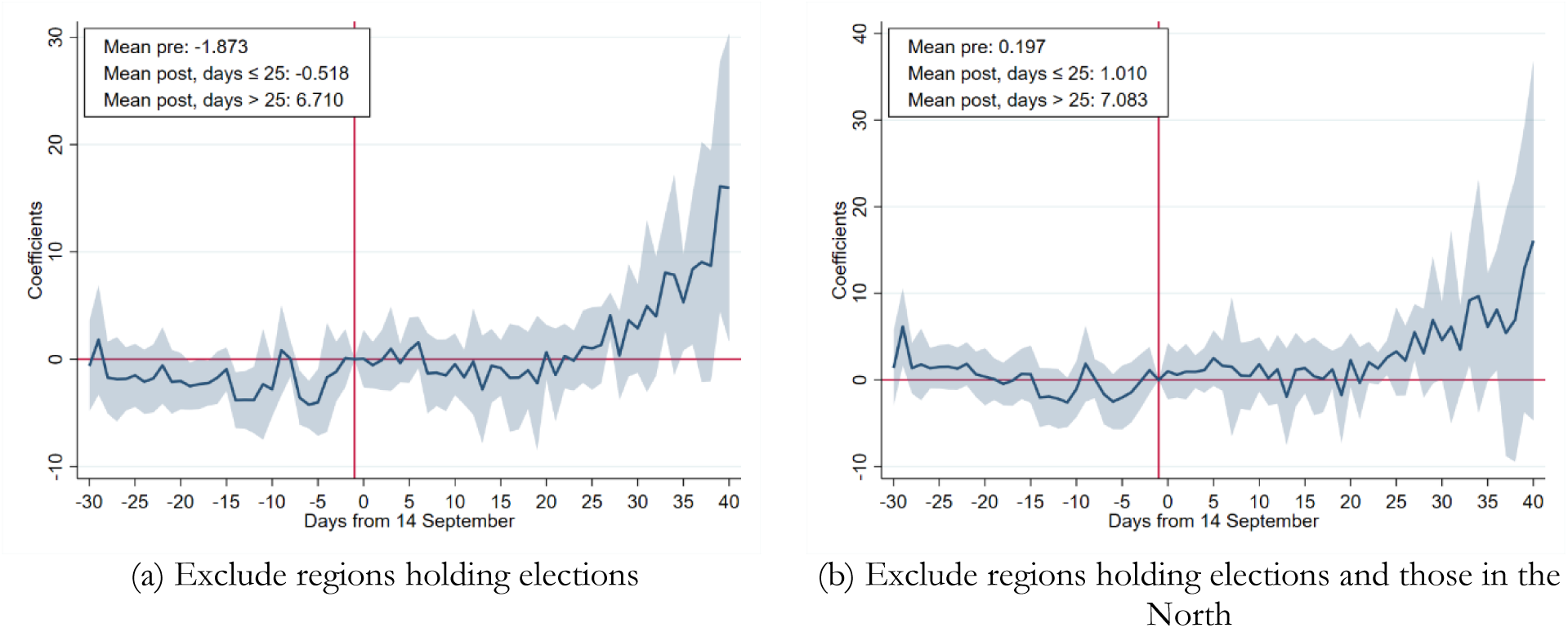
Difference-in-differences coefficients for daily cases, regions with no elections *Notes*. The figure reports estimates of dynamic coefficients *β*_*k*_ from equation (1), restricting the sample to regions not holding elections in panel (a) and, additionally, to regions in the Centre-South of the country in panel (b). Regions included in the treated group in panel (a) are: Emilia-Romagna, Lazio, Lombardia, Molise, Trento Province, Piemonte, Sicilia and Umbria; in panel (b): Lazio, Molise, Sicilia and Umbria. In both panels (a) and (b) Abruzzo, Basilicata, and Calabria are in the control group. Outcomes are values per 100,000 residents. The shaded areas are 95 percent confidence intervals obtained from cluster-robust standard errors at the region level.

**Figure A5.**
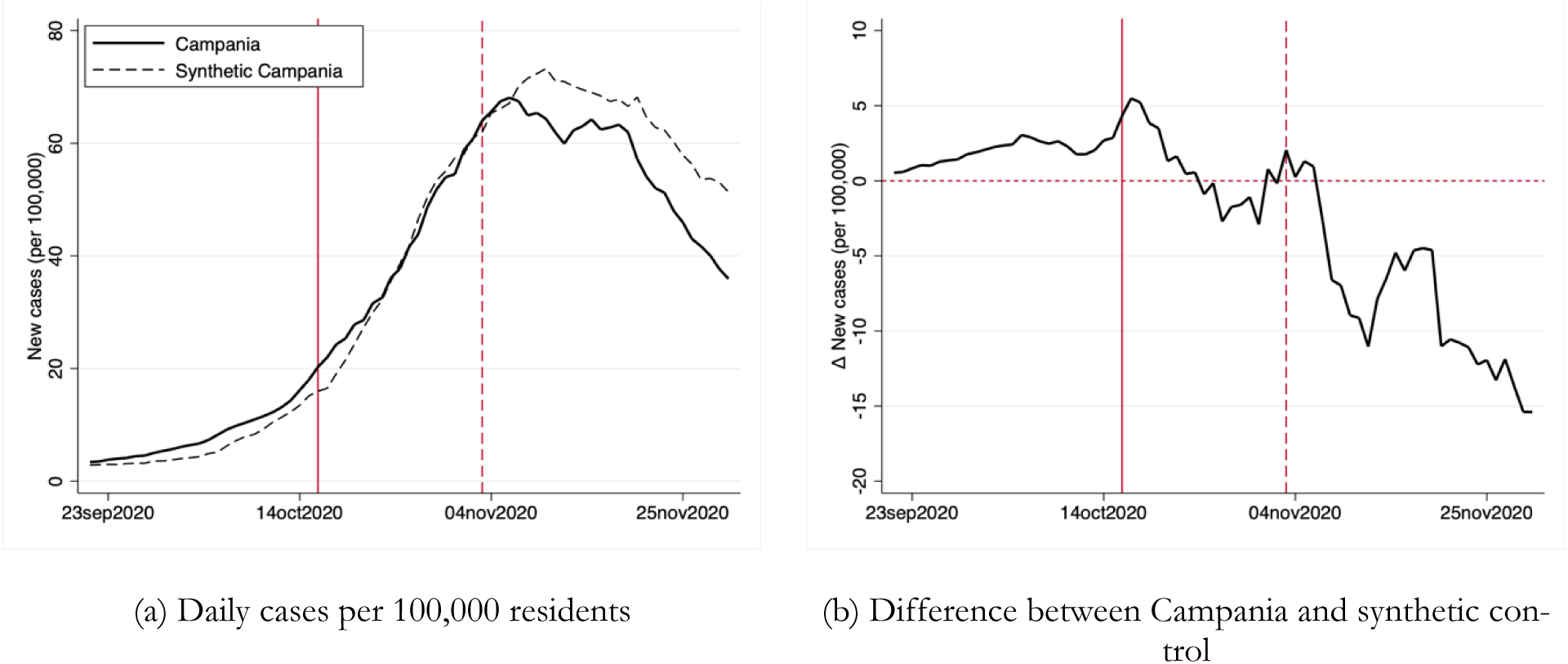
Evolution of new daily cases (7-day moving average) per 100,000 residents in Campania and in the synthetic control group, excluding Bolzano among regions in the control group *Notes*. The figure reports in panel (a) the evolution of daily cases in Campania (solid line) and in the synthetic control group (dashed line) between 21 September and 30 November. Panel (b) reports the difference between the two lines. The solid vertical line corresponds to 16 October, when schools were closed in Campania. The dashed vertical line corresponds to 3 November, when new nation-wide non pharmaceutical interventions were adopted by the central government. The sample excludes Bolzano in the control group.

**Figure A6.**
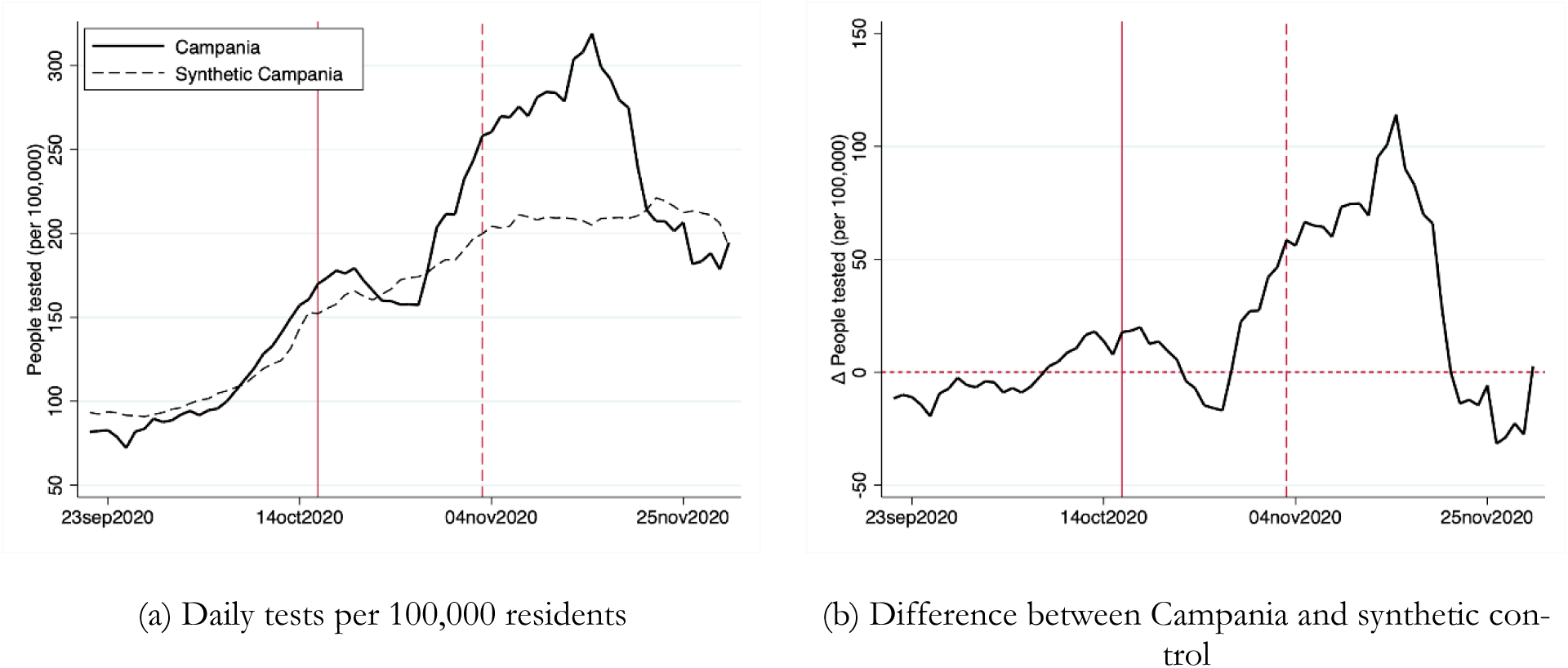
Evolution of new daily tests (7-day moving average) per 100,000 residents in Campania and in the synthetic control group *Notes*. The figure reports in panel (a) the evolution of daily tests in Campania (solid line) and in the synthetic control group (dashed line) between 21 September and 30 November. Panel (b) reports the difference between the two lines. The solid vertical line corresponds to 16 October, when schools were closed in Campania. The dashed vertical line corresponds to 3 November, when new nation-wide non pharmaceutical interventions were adopted by the central government.

**Figure A7.**
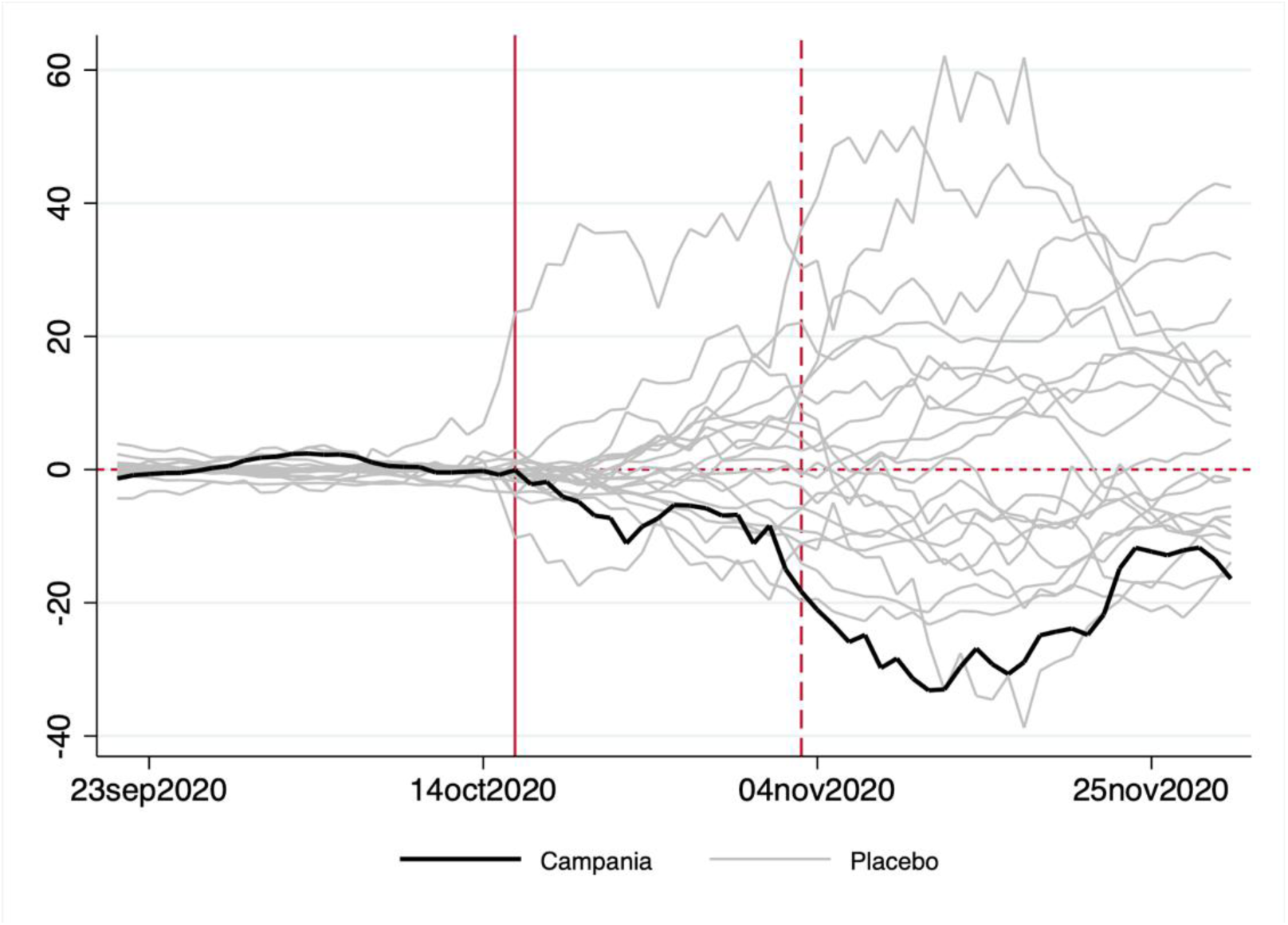
Distribution of synthetic control estimates using different regions as placebo units *Notes*. The figure reports the distribution of placebo estimates, obtained by computing the difference in new cases per 100,000 residents between a placebo treatment group and the synthetic control group. The placebo treatment group is one of the twenty Italian regions (eighteen regions plus two autonomous provinces, besides Campania). The black thick line is the true estimate using Campania as treated region. The solid vertical line corresponds to 16 October, when schools were closed in Campania. The dashed vertical line corresponds to 3 November, when new nation-wide non pharmaceutical interventions were adopted by the central government.

**Figure A8.**
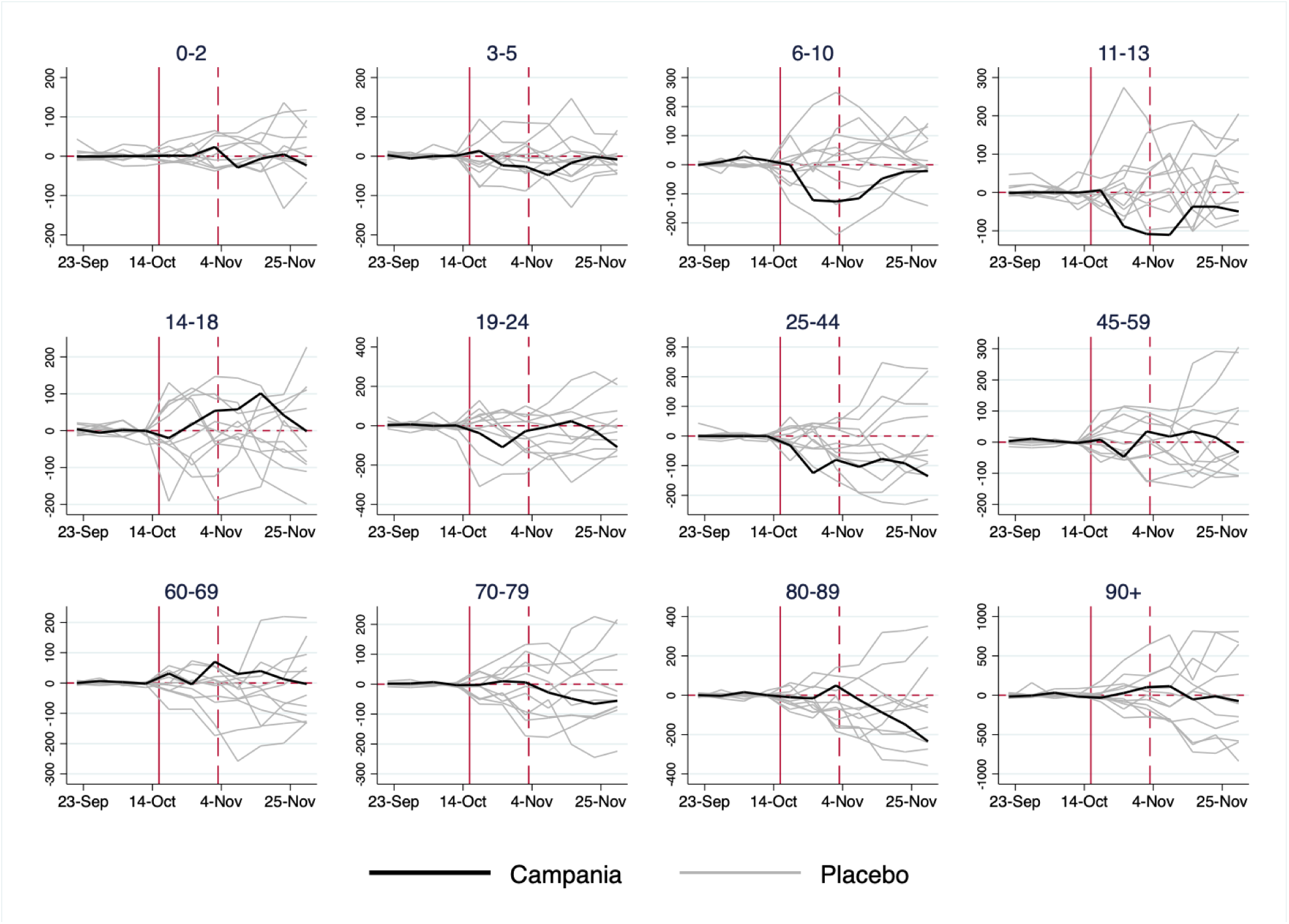
Distribution of synthetic control estimates by age group using different regions as placebo units *Notes*. The figure reports the distribution of placebo estimates, obtained by computing the difference in new cases per 100,000 residents between a placebo treatment group and the synthetic control group, for each age group. The placebo treatment group is one of the twelve Italian regions available in the dataset by age. The black thick line is the true estimate using Campania as treated region. The solid vertical line corresponds to 16 October, when schools were closed in Campania. The dashed vertical line corresponds to 3 November, when new nation-wide non pharmaceutical interventions were adopted by the central government.

**Figure A9.**
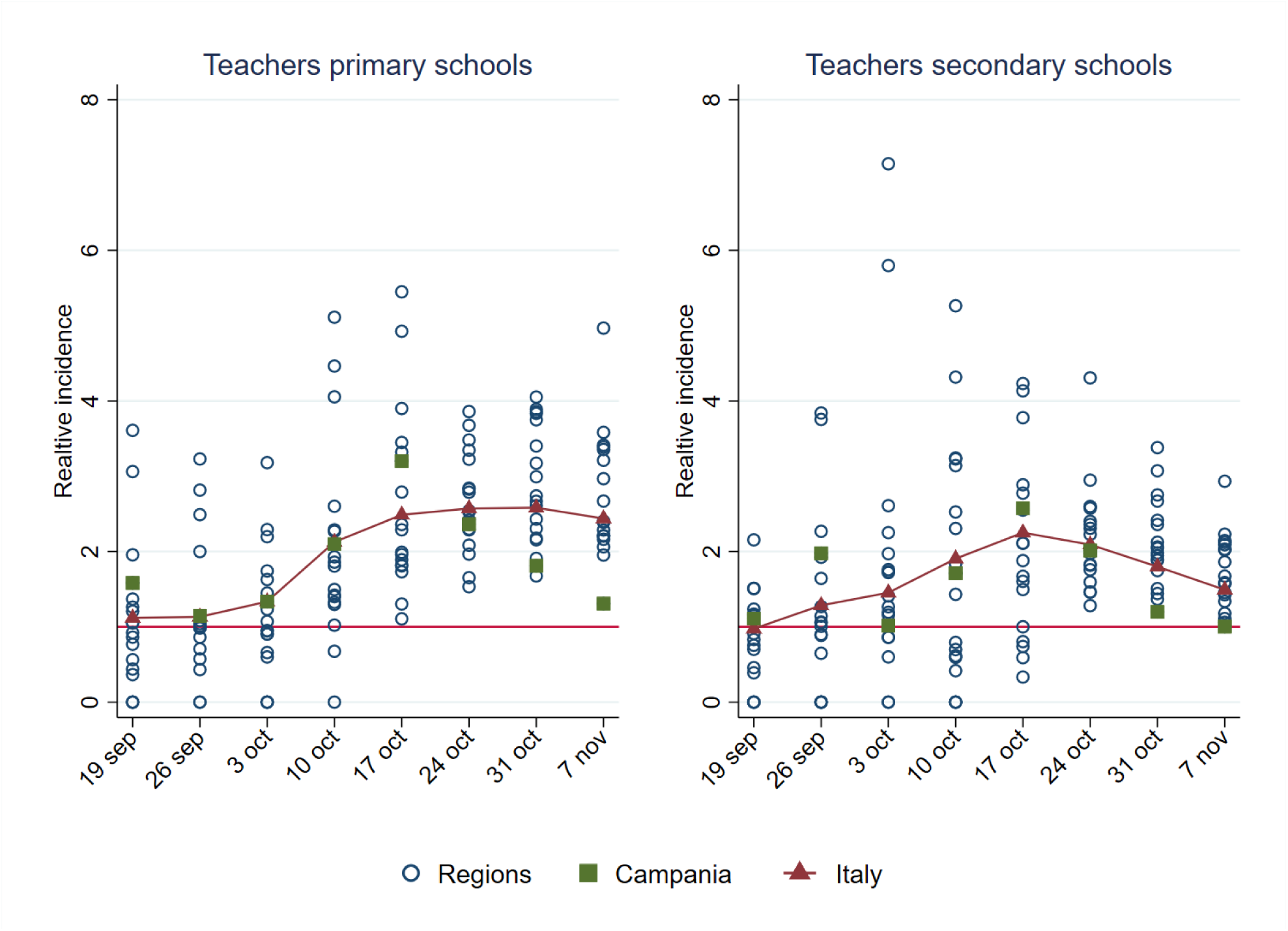
Incidence rate in teachers of primary and secondary schools relative to that in the general population in Italy and in each region between 19 September and 7 November *Notes*. The figure reports the relative incidence for teachers of primary and secondary schools, computed as the ratio of incidence for each group relative to incidence of the population. Source: author’s own elaboration based on data from the Ministry of Education and *Protezione Civile*.

**Figure A10.**
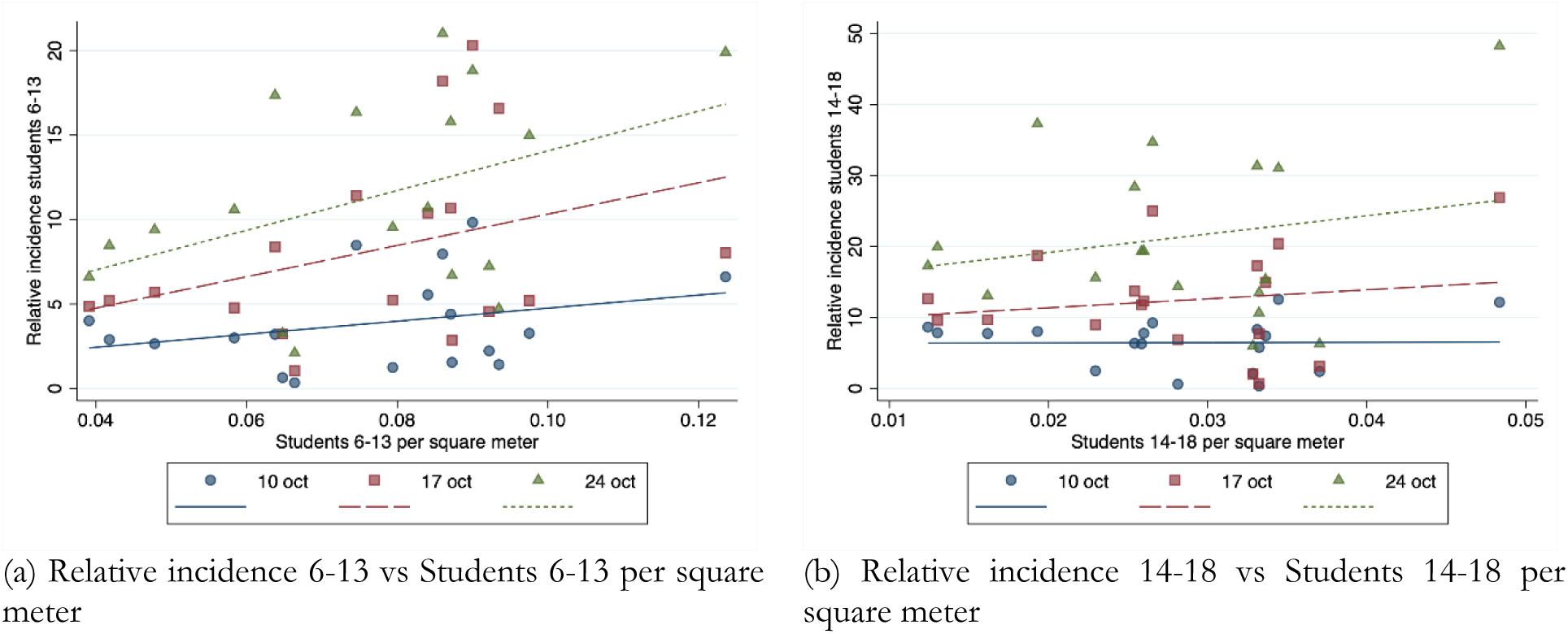
Regional-level scatter plots of the relative incidence among students vs the number of students per square meter, age group 6-13 (panel a) and 14-18 (panel b). *Notes*. The figure reports regional level scatter plots and linear fit of the relative incidence among students in the weeks of 10, 17 and 24 October against the number of students per square meter for the age groups 6-13 (panel a) and 14-18 (panel b).

**Table A1.** Synthetic control weights

|  | (1) | (2) | (3) | (4) | (5) | (6) | (7) | (8) | (9) | (10) | (11) | (12) | (13) |
| --- | --- | --- | --- | --- | --- | --- | --- | --- | --- | --- | --- | --- | --- |
|  | Age group |  |  |  |  |  |  |  |  |  |  |  |  |
| Region | All | 0-2 | 3-5 | 6-10 | 11-13 | 14-18 | 19-24 | 25-44 | 45-59 | 60-69 | 70-79 | 80-89 | 90+ |
| Abruzzo | 0 | - | - | - | - | - | - | - | - | - | - | - | - |
| Basilicata | 0 | - | - | - | - | - | - | - | - | - | - | - | - |
| Calabria | 0 | - | - | - | - | - | - | - | - | - | - | - | - |
| Emilia-Romagna | 0 | 0.598 | 0.264 | 0 | 0 | 0 | 0.324 | 0 | 0 | 0 | 0 | 0 | 0 |
| Friuli Venezia Giulia | 0 | 0 | 0 | 0 | 0 | 0 | 0 | 0 | 0 | 0 | 0 | 0 | 0 |
| Lazio | 0.295 | 0 | 0 | 0.636 | 0.644 | 0 | 0.400 | 0.576 | 0.884 | 0.969 | 0 | 0 | 0 |
| Liguria | 0 | - | - | - | - | - | - | - | - | - | - | - | - |
| Lombardia | 0 | 0.150 | 0.319 | 0.364 | 0 | 0 | 0.084 | 0.141 | 0 | 0 | 0 | 0.109 | 0.025 |
| Marche | 0 | 0 | 0 | 0 | 0 | 0 | 0 | 0 | 0 | 0 | 0 | 0 | 0 |
| Molise | 0 | - | - | - | - | - | - | - | - | - | - | - | - |
| P.A. Bolzano | 0.662 | - | - | - | - | - | - | - | - | - | - | - | - |
| P.A. Trento | 0 | 0 | 0 | 0 | 0 | 0 | 0 | 0 | 0 | 0 | 0 | 0 | 0 |
| Piemonte | 0 | 0 | 0 | 0 | 0 | 0.135 | 0.036 | 0.021 | 0.014 | 0.031 | 0.114 | 0 | 0 |
| Puglia | 0 | 0 | 0 | 0 | 0 | 0 | 0 | 0 | 0 | 0 | 0 | 0 | 0 |
| Sardegna | 0 | - | - | - | - | - | - | - | - | - | - | - | - |
| Sicilia | 0 | 0 | 0 | 0 | 0.131 | 0.725 | 0 | 0 | 0.101 | 0 | 0.886 | 0.277 | 0.975 |
| Toscana | 0 | 0.252 | 0.326 | 0 | 0.098 | 0 | 0 | 0 | 0 | 0 | 0 | 0 | 0 |
| Umbria | 0 | 0 | 0 | 0 | 0 | 0 | 0 | 0 | 0 | 0 | 0 | 0 | 0 |
| Valle d'Aosta | 0.043 | - | - | - | - | - | - | - | - | - | - | - | - |
| Veneto | 0 | 0 | 0.092 | 0 | 0.127 | 0.140 | 0.157 | 0.262 | 0 | 0 | 0 | 0.613 | 0 |

*Notes.* The table reports the weights used to compute the synthetic control group for each outcome. Column (1) reports the weights used to analyze the daily number of cases per 100,000 residents. Columns (2) to (13) report the weights for incidence rates for different age groups. In the latter case, the sample of regions is limited to 13, because of data availability.

**Table A2.** Association between the number of students per square meter and the relative incidence among students, 6-13 and 14-18 years old, OLS estimates

| *Relative incidence in week starting:* | (1) | (2) | (3) | (4) | (5) | (6) |
| --- | --- | --- | --- | --- | --- | --- |
|  | 10 October |  | 17 October |  | 24 October |  |
| **[a] Students 6-13 years old** |  |  |  |  |  |  |
| Students 6-13 per square meter | 38.67*<br>(19.14) | 38.72<br>(32.41) | 92.68*<br>(45.70) | 122.21<br>(69.55) | 117.50**<br>(41.48) | 149.52**<br>(51.77) |
| GDP per capita |  | 0.17*<br>(0.09) |  | 0.24<br>(0.17) |  | 0.55***<br>(0.10) |
| PM10 |  | -0.13<br>(0.24) |  | 0.26<br>(0.61) |  | 0.16<br>(0.34) |
| Population density |  | 0.01<br>(0.01) |  | -0.01<br>(0.03) |  | 0.00<br>(0.01) |
| Constant | 0.90<br>(1.34) | -1.69<br>(6.20) | 1.06<br>(2.80) | -11.97<br>(12.71) | 2.32<br>(3.08) | -19.38*<br>(9.58) |
| Standardized effect | 0.83 | 0.84 | 2.00 | 2.64 | 2.54 | 3.23 |
| Observations | 18 | 18 | 18 | 18 | 18 | 18 |
| R-squared | 0.09 | 0.32 | 0.13 | 0.23 | 0.19 | 0.72 |
| **[b] Students 14-18 years old** |  |  |  |  |  |  |
| Students 14-18 per square meter | 4.17<br>(108.12) | -23.75<br>(114.21) | 126.57<br>(222.20) | 275.37<br>(252.43) | 258.35<br>(396.81) | 576.14*<br>(285.43) |
| GDP per capita |  | 0.25**<br>(0.10) |  | 0.68**<br>(0.24) |  | 1.35***<br>(0.33) |
| PM10 |  | -0.31<br>(0.33) |  | 0.14<br>(1.01) |  | 0.18<br>(1.01) |
| Population density |  | 0.02<br>(0.01) |  | 0.01<br>(0.04) |  | -0.00<br>(0.04) |
| Constant | 6.35**<br>(2.65) | 3.73<br>(10.09) | 8.84<br>(5.36) | -19.08<br>(27.35) | 14.01<br>(9.95) | -35.57<br>(28.77) |
| Standardized effect | 0.04 | -0.22 | 1.15 | 2.50 | 2.34 | 5.23 |
| Observations | 18 | 18 | 18 | 18 | 18 | 18 |
| R-squared | 0.00 | 0.54 | 0.02 | 0.59 | 0.04 | 0.71 |

*Notes.* The table reports region-level estimates from OLS regressions of the relative incidence among students on the number of students per square meter between 6 and 13 years (panel a) and between 14 and 18 years old (panel b). The sample includes all Italian regions except Valle d'Aosta and Trentino Alto Adige, for which data on incidence rates is not available. The dependent variables in columns 1 and 2 is the age-specific incidence rate in the week of 10 October; in columns 3 and 4, it is the age-specific incidence rate in the week of 17 October; in columns 5 and 6 it is the age-specific incidence rate in the week of 24 October. See text for details on control variables included in columns 2, 4, and 6. The "standardized effect" equals the coefficients on the main explanatory variable (students per square meter) multiplied by its standard deviation. Robust standard errors in parentheses. Significance levels: \* $p < 0.1$ ; \*\* $p < 0.05$ ; \*\*\* $p < 0.01$ .

## Footnotes

1 The school closure tracker is available online at https://ourworldindata.org/covid-school-workplace-closures. Last access: July 14, 2021.

2 Differences between studies can also emerge in the presence of different research designs and sample selections. In particular, Gandini et al. (2021) and I both examine Italy as a case study, but they conclude that schools did not contribute to the spread of the virus. In particular, using the same data source from the Ministry of Education, they conclude that incidence among students and teachers is lower that than in the general population. I find an opposite result using different time periods than theirs.

3 See Werner and Woessmann (2021) for a review of the evidence on the effects of the pandemic on children’s cognitive and socio-emotional developments, and Stantcheva (2022) for a review of the literature on the comprehensive effects of the pandemic on inequalities, comprising that stemming from school closures.

4 There are 21 regions (19 regions and two autonomous provinces) and 107 provinces in Italy.

5 Data on incidence rates by age group are not available before that date. Therefore, they cannot be used to analyze the impact of school re-openings in September, as they are missing for the pre-treatment period.

6 The data can be downloaded from the webpage of the article in the <u>Supplementary materials section</u>. Last access 19 July 2021.

7 Portale Unico dei Dati della Scuola, available at https://dati.istruzione.it/opendata/.

8 Specifically, each point in the graph is the predicted value from a regression of mobility on week dummies, a treatment dummy (equal to one for early opening regions) and the interaction of both. 95 percent confidence intervals are recovered from standard errors obtained via the delta method.

9 I do not include mobility towards transit stations among controls, as it is directly affected by school openings. Results – available upon requests – are nonetheless similar if I include it among controls.

10 For the age groups 0-2, I only use the three adjacent older group (3-5, 6-10, 11-13). For the age group over 90, I include the three adjacent younger groups (60-69, 70-79, 80-89).

11 Using the notation of equation (1), I am computing 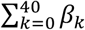.

12 In contrast, Armillei (2021) suggests that local elections had a minor role in contributing to the dynamics of the epidemic.

13 Therefore, in both panels (a) and (b), the control group comprises three regions: Abruzzo, Basilicata and Calabria. The treatment group of early opening regions includes Emilia-Romagna, Lazio, Lombardia, Molise, Trento Autonomous Province, Piemonte, Sicilia and Umbria in panel (a), and Lazio, Molise, Sicilia and Umbria in panel (b).

14 I use moving averages to smooth lines and avoid fluctuations due to idiosyncratic factors affecting daily cases when looking at a single region.

15 Almost two-thirds of the synthetic control group in the aggregate analysis is accounted by the province of Bolzano. I construct a new synthetic control group excluding Bolzano from the pool of candidate regions in the control group and report the resulting estimates for the evolution of daily cases in Figure A5. The evolution of daily cases in Campania before school closures does not match perfectly that in the control group, but there is a clear declining trend after 16 October. Note that in this case the control group comprises Toscana and Sardinia, with almost equal weights.

16 Source: EEA Airbase database: https://www.eea.europa.eu/data-and-maps/data/aqereporting-9.

17 One additional potential mechanism behind cross-country differences, which cannot be explored empirically with the data at hand, is the presence of mask mandates for children in schools. Chernozhukov et al. (2021) show, for example, that in US counties with mask mandates, COVID-19 cases increased less following school openings.

## References

Abadie, Alberto, Alexis Diamond, and Jens Hainmueller. 2010. ‘Synthetic Control Methods for Comparative Case Studies: Estimating the Effect of California’s Tobacco Control Program’. Journal of the American Statistical Association 105 (490): 493–505.

Adda, Jérôme. 2016. ‘Economic Activity and the Spread of Viral Diseases: Evidence from High Frequency Data’. The Quarterly Journal of Economics 131 (2): 891–941.

Agostinelli, Francesco, Matthias Doepke, Giuseppe Sorrenti, and Fabrizio Zilibotti. 2022. ‘When the Great Equalizer Shuts Down: Schools, Peers, and Parents in Pandemic Times’. Journal of Public Economics 206: 104574.

Alon, Titan, Matthias Doepke, Jane Olmstead-Rumsey, and Michèle Tertilt. 2020. ‘The Impact of COVID-19 on Gender Equality’. National Bureau of Economic Research Working Paper Series No. 26947.

Amodio, Emanuele, Michele Battisti, Andros Kourtellos, Giuseppe Maggio, and Carmelo Massimo Maida. 2021. ‘Schools Opening and COVID-19 Diffusion: Evidence from Geolocalized Microdata’. European Economic Review 143: 104003.

Armillei, Francesco. 2021. ‘Rinvio Delle Amministrative: Forse Un Eccesso Di Prudenza’. Lavoce.Info, 6 April 2021. https://www.lavoce.info/archives/73349/rinvio-delle-amministrative-forse-un-eccesso-di-prudenza/.

Bravata, Dena, Jonathan H Cantor, Neeraj Sood, and Christopher M Whaley. 2021. ‘Back to School: The Effect of School Visits During COVID-19 on COVID-19 Transmission’. National Bureau of Economic Research Working Paper Series No. 28645.

Chernozhukov, Victor, Hiroyuki Kasahara, and Paul Schrimpf. 2021. ‘The Association of Opening K-12 Schools and Colleges with the Spread of COVID-19 in the United States: County-Level Panel Data Analysis’. MedRxiv, January, 2021.02.20.21252131.

Cipullo, Davide, and Marco Le Moglie. 2022. ‘To Vote, or Not to Vote: On the Epidemiological Impact of Electoral Campaigns at the Time of COVID-19’. European Journal of Political Economy 72, 102118.

Contini, Dalit, Maria Laura Di Tommaso, Caterina Muratori, Daniela Piazzalunga, and Lucia Schiavon. 2022. ‘Who Lost the Most? Mathematics Achievement during the COVID-19 Pandemic’. The B.E. Journal of Economic Analysis & Policy 22 (2): 399–408.

Courtemanche, Charles J, Anh H Le, Aaron Yelowitz, and Ron Zimmer. 2021. ‘School Reopenings, Mobility, and COVID-19 Spread: Evidence from Texas’. National Bureau of Economic Research Working Paper Series No. 28753.

Engzell, Per, Arun Frey, and Mark D Verhagen. 2021. ‘Learning Loss Due to School Closures during the COVID-19 Pandemic’. Proceedings of the National Academy of Sciences 118 (17): e2022376118.

European Parliament, Directorate-General for Internal Policies of the Union, L Smit, M Koopmans, B Brunekreef, G Hoek, G Downward, et al. 2021. Air Pollution and COVID-19 : Including Elements of Air Pollution in Rural Areas, Indoor Air Pollution, Vulnerability and Resilience Aspects of Our Society against Respiratory Disease, Social Inequality Stemming from Air Pollution.

Fuchs-Schündeln, Nicola, Dirk Krueger, Alexander Ludwig, and Irina Popova. 2022. ‘The Long-Term Distributional and Welfare Effects of COVID-19 School Closures’. The Economic Journal 132(645): 1647–1683

Gandini, Sara, Maurizio Rainisio, Maria Luisa Iannuzzo, Federica Bellerba, Francesco Cecconi, and Luca Scorrano. 2021. ‘A Cross-Sectional and Prospective Cohort Study of the Role of Schools in the SARS-CoV-2 Second Wave in Italy’. The Lancet Regional Health – Europe 5.

Goldhaber, Dan, Scott A Imberman, Katharine O Strunk, Bryant Hopkins, Nate Brown, Erica Harbatkin, and Tara Kilbride. 2021. ‘To What Extent Does In-Person Schooling Contribute to the Spread of COVID-19? Evidence from Michigan and Washington’. National Bureau of Economic Research Working Paper Series No. 28455.

Halloran, Clare, Rebecca Jack, James C Okun, and Emily Oster. 2021. ‘Pandemic Schooling Mode and Student Test Scores: Evidence from US States’. National Bureau of Economic Research Working Paper Series No. 29497.

Head, Jennifer R, Kristin L Andrejko, Qu Cheng, Philip A Collender, Sophie Phillips, Anna Boser, Alexandra K Heaney, et al. 2022. ‘School Closures Reduced Social Mixing of Children during COVID-19 with Implications for Transmission Risk and School Reopening Policies’. Journal of The Royal Society Interface 18 (177): 20200970.

Isphording, Ingo E, Marc Lipfert, and Nico Pestel. 2021. ‘Does Re-Opening Schools Contribute to the Spread of SARS-CoV-2? Evidence from Staggered Summer Breaks in Germany’. Journal of Public Economics 198: 104426.

Jackson, Charlotte, Emilia Vynnycky, Jeremy Hawker, Babatunde Olowokure, and Punam Mangtani. 2013. ‘School Closures and Influenza: Systematic Review of Epidemiological Studies’. BMJ Open 3 (2): e002149.

Lee, Joyce. 2020. ‘Mental Health Effects of School Closures during COVID-19’. The Lancet Child & Adolescent Health 4 (6): 421.

Parolin, Zachary, and Emma K Lee. 2021. ‘Large Socio-Economic, Geographic and Demographic Disparities Exist in Exposure to School Closures’. Nature Human Behaviour 5 (4): 522–28.

Stantcheva, Stefanie. 2022. ‘Inequalities in the Times of a Pandemic’. National Bureau of Economic Research Working Paper Series No. 29657.

von Bismarck-Osten, Clara, Kirill Borusyak, and Uta Schonberg. 2022. ‘The Role of Schools in Transmission of the SARS-CoV-2 Virus: Quasi-Experimental Evidence from Germany’. Economic Policy 37(109): 87–130.

Werner, Katharina, and Ludger Woessmann. 2021. ‘Will the COVID-19 Pandemic Leave a Lasting Legacy in Children’s Skill Development?’ CESifo Forum, CESifo Forum, 22 (06): 33–40.

